# Genetically personalised organ-specific metabolic models in health and disease

**DOI:** 10.1101/2022.03.25.22272958

**Authors:** Carles Foguet, Yu Xu, Scott C. Ritchie, Samuel A. Lambert, Elodie Persyn, Artika P. Nath, Emma E. Davenport, David J. Roberts, Dirk S. Paul, Emanuele Di Angelantonio, John Danesh, Adam S. Butterworth, Christopher Yau, Michael Inouye

**Author notes:** Corresponding authors (CF) & (MI).

## Abstract

Understanding how genetic variants influence disease risk and complex traits (variant-to-function) is one of the major challenges in human genetics. Here we present a model-driven framework to leverage human genome-scale metabolic networks to define how genetic variants affect biochemical reaction fluxes across major human tissues, including skeletal muscle, adipose, liver, brain and heart. As proof of concept, we build personalised organ-specific metabolic flux models for 524,615 individuals of the INTERVAL and UK Biobank cohorts and perform a fluxome-wide association study (FWAS) to identify 4,411 associations between personalised flux values and the concentration of metabolites in blood. Furthermore, we apply FWAS to identify 97 metabolic fluxes associated with the risk of developing coronary artery disease, many of which are linked to processes previously described to play in role in the disease. Our work demonstrates that genetically personalised metabolic models can elucidate the downstream effects of genetic variants on biochemical reactions involved in common human diseases.

## Introduction

Genome-wide association studies (GWAS) have identified more than 50,000 genetic variants associated with complex traits or diseases^1^. While the contribution of individual variants to a given phenotype is generally small, the effect of multiple genetic variants can be aggregated into polygenic scores (PGS), which are highly predictive of disease incidence and enhance existing risk models^2–4^. However, while GWAS and PGS can be useful for risk stratification^5–7^, the mechanisms by which genetic variants influence disease risk, i.e. variant to function (V2F), remain largely unsolved. Addressing V2F is a major challenge in human genetics and has the potential to unveil many new therapeutic targets^6,8,9^.

An approach to address the V2F challenge is to quantify how genetic variation causes disease through the regulation of molecular traits. To this end, genetic variants affecting gene expression are identified and subsequently aggregated into models that can impute the abundances of transcripts and proteins^10,11^. For example, PredictDB is a database that offers a collection of linear models to impute transcript levels in specific organs of the human body^12^. PredictDB models were trained in the GTEx dataset, which contains genotype profiling and tissue-specific transcript abundance from post-mortem donors^13^. Imputed transcript or protein levels can be used to perform transcriptome-wide or proteome-wide association analyses, respectively, to identify gene products associated with disease^10,14^. Alternately, PGSs for disease can be used to identify proteins and other gene products which may disrupt polygenic risk^15^. However, transcripts and proteins do not exert their effects in isolation but in highly connected and complex biological networks. Indeed, previous studies have shown the merit of analysing genetic variation in the context of gene co-expression and gene interaction networks to characterise how the effects of genetic variants contribute to complex traits or disease by propagating through biological networks^16–19^

Metabolism is one of the most prominent biological networks and a comparatively tractable experimental setting in which to address the V2F challenge. Essentially, metabolism is a set of interconnected chemical reactions and transport processes occurring in a highly ordered, regulated and coordinated manner across multiple organs in the human body^20^. The metabolic phenotype of a given organ is defined by both metabolite concentrations and metabolic fluxes (i.e. the rates at which substrates are converted to products through reactions) and emerges from the complex interaction of metabolites, enzymes and transmembrane carriers^21,22^. Metabolite concentrations offer a static snapshot of metabolite distributions, whereas metabolic fluxes provide a map of metabolite traffic through metabolic pathways^23^.

Genome-scale metabolic models (GSMMs), mathematical representations of the metabolic reaction network arising from the human genome^24,25^, simulate steady-state metabolic fluxes by formulating network stoichiometry as sets of linear equations and directionality constraints^26^. GSMMs have emerged as a useful approach to integrate transcriptomics, proteomics, and metabolomics to characterise metabolic flux maps^27,28^. For example, proteomics, metabolomics and physiological data have been used to build human organ-specific GSMMs^29^. Similarly, there is increasing interest to integrate individual measures to build personalised GSMMs that reflect the specific metabolic phenotype in each individual, thus facilitating personalised medicine^29–33^.

Since gene expression is highly heritable^10,12^, it may be feasible to leverage human genome-scale metabolic networks to analyse the system-wide effects of genetic variants on metabolism and build genetically personalised GSMMs. To this end, we present a framework where transcript levels imputed from genetic data can be used to simulate personalised and organ-specific, genome-scale flux maps using the quadratic metabolic transformation algorithm (qMTA). Such flux maps provide genetically personalised metabolic models at a genome scale for each tissue. As proof of concept, we build personalised organ-specific flux maps for over 520,000 individuals across the INTERVAL^34,35^ and UK Biobank (UKB)^36^ cohorts, then perform a fluxome-wide association study (FWAS) to test the association between organ-specific flux values and directly measured blood metabolite levels. Finally, we apply FWAS to identify fluxes associated with coronary artery disease (CAD), thus demonstrating the potential of genome-scale flux maps for V2F by elucidating intermediary biochemical reactions between genetic variation and common disease.

## Results

### A computational framework for genetically personalised organ-specific GSMMs

We developed a framework for building personalised organ-specific flux maps from genotype data (**Fig. 1; Methods**). The first step is to compute a reference flux distribution for each organ under consideration. This is achieved by extracting the organ-specific subnetwork from the Harvey/Harvetta multiorgan model^29^, defining organ-specific metabolic functions that must be fulfilled (e.g. synthesis of neurotransmitters in the brain), obtaining average organ transcript abundance from GTEx^13^ and using the transcript abundance as an input for the GIM3E algorithm^37^. GIM3E is an algorithm that, subject to maintaining the organ-specific metabolic functions, seeks to minimise the overall flux through the network using transcript abundance data to give each reaction a minimisation weight inversely proportional to the expression of the enzymes catalysing it. Subsequently, flux sampling^38^ is applied to identify a representative solution within the solution space and within 99% of the GIM3E optimal solution. The resulting set of flux values, termed reference flux distribution, is both enzymatically efficient and consistent with the average transcript abundance in each organ (**Fig. S1**). The flux distribution can be assumed to represent the average metabolic state of each modelled organ in the sampled population.

**Fig. 1:**
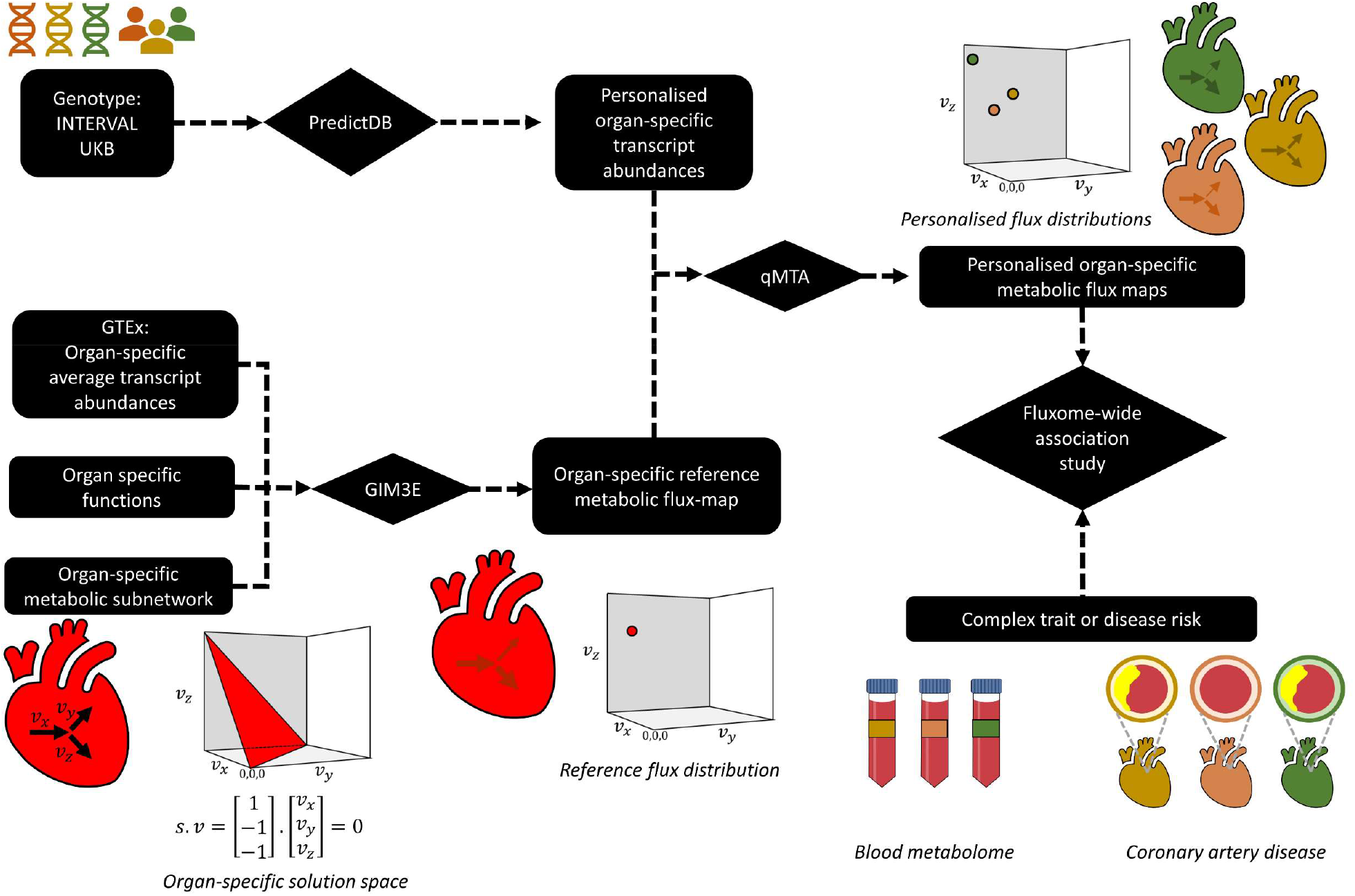
Framework for computing organ-specific personalised genome-scale flux maps from genotype data and performing a fluxome-wide association study (FWAS). First, a reference flux map is computed for each organ using the GIM3E algorithm to integrate average transcript abundances and organ-specific metabolic functions into an organ-specific metabolic subnetwork. In parallel, personalised organ-specific transcript abundances are imputed from genotype data of the INTERVAL and UKB cohorts using the models from PredictDB. Next, the quadratic metabolic transformation algorithm (qMTA) is used to integrate the organ-specific transcript abundances and reference flux distribution and compute personalised organ-specific metabolic flux maps. The resulting flux maps can be used to perform FWAS to complex traits or diseases such as blood metabolic features or coronary artery disease. A hypothetic representation of an organ-specific solution space, reference flux distribution, and personalised flux distributions is shown for a reaction network with three fluxes (*v*_*x*_, *v*_*y*_ and *v*_*z*_).

Subsequently, models from PredictDB^12^ are used to impute organ-specific transcript abundances from genotype data. The resulting imputed transcript data are mapped to reactions in the organ-specific subnetworks derived from the Harvey/Harvetta multiorgan model^29^ as putative reaction activity fold changes relative to the average organ-specific transcript expression in GTEx^13^. The imputed reaction activity fold changes and the reference flux distributions are then utilised by the qMTA to compute genetically personalised organ-specific flux maps (**Fig. S2**). Briefly, qMTA finds the flux distributions (i.e., sets of flux values) most consistent with the putative reaction activity fold changes in each individual (**Methods**).

### Building flux maps for >520,000 individuals

Using the above framework, we built personalised organ-specific flux maps for 37,220 and 487,395 individuals from the INTERVAL^34,35^ and UKB^36^ cohorts, respectively. Personalised models were generated for skeletal muscle, adipose tissue, liver, brain, and heart, which together account for roughly 66% of body weight in an average adult^39^. Overall, 16,058 reaction flux values were computed for each individual. Metabolic fluxes “flow” through pathways where the product of one reaction is the substrate of successive reactions; thus, many of the flux values computed in each individual will have inherent dependencies (**Fig. S3**). As such, from the 16,058 reaction flux values, we selected a subset of 4,418 flux values without strong pairwise correlations (ρ<0.9) for further analysis (**Fig. S4; Methods**).

Principal component analysis of the personalised organ-specific flux values for individuals of INTERVAL and UKB showed the underlying structure in the data (**Fig. S5**). Fluxes with the greatest loadings on top principal components (PCs) tended to be related to the known metabolism of each organ (**Table S1**). For example, in the liver, fluxes through reactions and transport processes of amino acid and lipid metabolism exhibited large loadings along the first five PCs. These included reactions linked to urea and acetaldehyde, which are predominantly metabolised in the liver^20,40^. Similarly, in skeletal muscle and heart, fluxes through transport processes of amino acid metabolism and reactions of fatty acid metabolism also had large PC loadings. However, in the heart, PC4 was also strongly associated with the flux through several reactions linked to the antioxidant defence system, which is associated with heart failure^41^. In the brain, PC1, PC2 and PC5 were strongly related to fluxes through reactions and transport processes of amino acid metabolism, including reactions linked to neurotransmitters such as dopamine, serotonin, or norepinephrine. However, PC3 was also strongly associated with fluxes linked to glycolipid metabolism, including gangliosides, a class of sialylated glycosphingolipids that predominate in nerve cells^42^. Finally, in adipose tissue, most PCs were associated with fluxes linked with lipid metabolism, such as the malic enzyme, which has a key role in *de novo* fatty acid synthesis^43^.

### Fluxome-wide association study for blood metabolites

We next validated that genetically personalised GSMMs could generate reliable and meaningful flux predictions across cohorts. As phenotypes, organ-specific flux maps are expected to lead to distinct profiles in the blood metabolome. To demonstrate this, we performed an association analysis by regressing measured blood metabolic features against personalised flux values in the INTERVAL^34,35^ and UKB^36^ cohorts (**Fig. S2**; **Methods**). The blood metabolome for INTERVAL comprised both Nightingale Health NMR assays (N= 37,720 participants) and Metabolon HD4 mass spec assays (N=8,115 participants)^44^. For UKB, Nightingale Health NMR was performed for 117,161 participants^45^.

For INTERVAL, an FDR-adjusted significance threshold of P<1.0×10^−6^ was applied to control for all tested pairs (**Methods**). We identified 4,411 significant associations between flux values and blood metabolic features in total, of which 1,406 were for the Nightingale platform and 3,005 for Metabolon (**Table S2, Fig. 2A-2B**). Consistent with the role of liver in whole-body metabolic homeostasis^20^, liver was the organ with the most associations (1,588), followed by heart (1,098), skeletal muscle (792), brain (536) and adipose tissue (397) (**Fig. 2C, Table S2**).

**Fig. 2:**
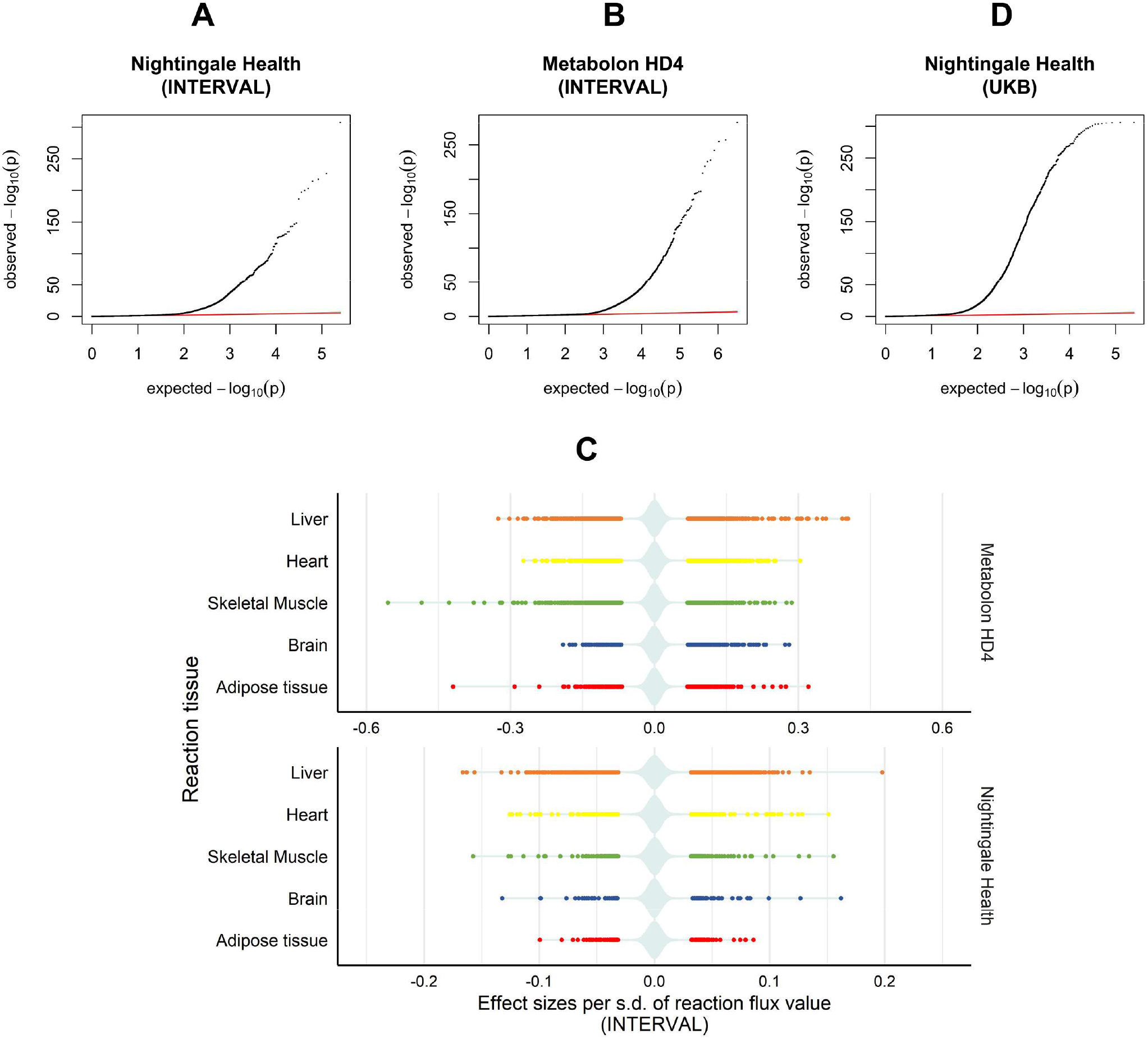
Fluxome-wide association study (FWAS) between genetically personalised flux maps and blood metabolic features. (**A**),(**B**),(**D**) quantile-quantile (QQ) plot of the observed P values for associations between flux values and blood metabolic features measured with the Nightingale Health platform in INTERVAL(**A**), Metabolon HD4 platform in INTERVAL(**B**) and Nightingale Health platform in UKB(**D**). (**C**) Plot of statistically significant flux effect sizes per organ to blood metabolic features measured with either the Nightingale Health or Metabolon HD4 assay in INTERVAL. A violin plot, coloured in pale azure, shows the distribution of both significant and non-significant effect sizes.

We externally validated the INTERVAL flux associations with Nightingale metabolites using UKB (**Fig. 2D)**. We found 83% of the INTERVAL associations replicated in UKB with FDR-adjusted significance threshold of P<1.0×10^−6^ and consistent direction of the effect sizes. Effect sizes were themselves highly correlated (ρ=0.82) between INTERVAL and UKB (**Fig. S6**).

### Fluxome associations by metabolic feature class and reaction subsystem

The 4,411 significant associations comprised 224 unique blood metabolic features and 649 unique organ-specific metabolic fluxes. Consistent with the coverage of the Nightingale Health and Metabolon HD4 platforms, we found that most of these blood metabolic features were lipid-related (**Fig. 3A, Table S2)**. Even so, Nightingale Health measured lipoprotein particles were enriched (**Methods**) in the associations of liver fluxes relative to all profiled metabolic features, consistent with the role of the liver in lipoprotein homoestasis^20^. Glycerides and phospholipids were enriched in associations across all organs relative to all features profiled with the Metabolon HD4 assay, suggesting an association with core reactions (i.e. active in all modelled organs). Reflecting the role of adipose tissue in steroid hormone metabolism^46^, adipose tissue was enriched in associations to steroids.

**Fig. 3:**
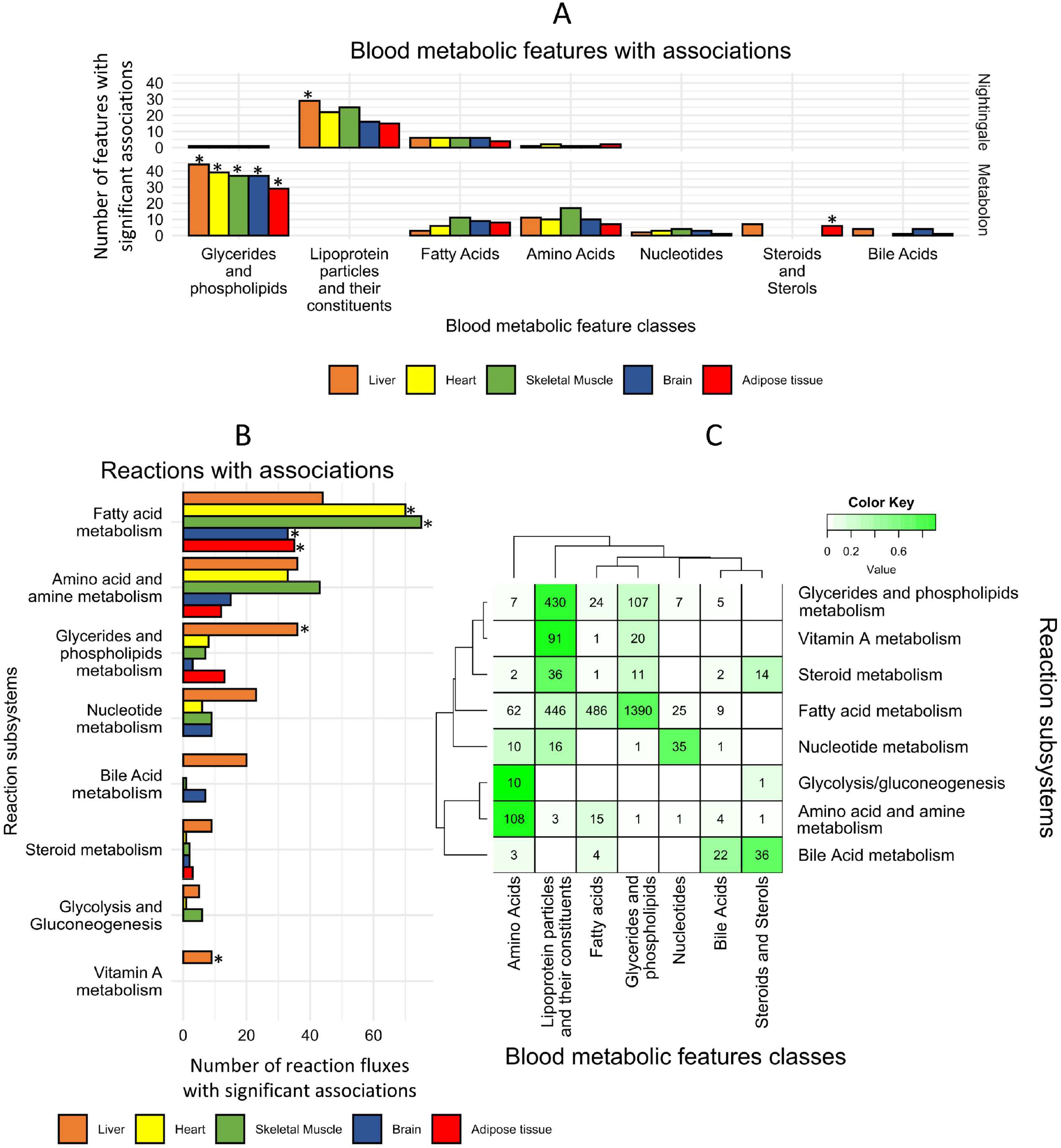
Characterisation of the significant associations between blood metabolic features and metabolic fluxes. (**A**) Classes of the blood metabolic features with one or more significant associations to the fluxome. *denotes classes that are significantly enriched (Fisher’s exact test, FDR-adjusted P value<0.05). Unannotated features and classes with few features are omitted for clarity. (**B**) Subsystems of the reactions whose flux values are significantly associated with one or more blood metabolic features. *denotes subsystems that are significantly enriched (Fisher’s exact test, FDR-adjusted P-value<0.05). Unannotated reactions and subsystems with few features are omitted for clarity. (**C**) Heatmap of the intersection between blood metabolic feature classes and reaction subsystem in significant associations. Numbers at each intersection denote the number of significant associations between reaction fluxes of a given subsystem and blood metabolic features of a given class. The colour key denotes the fraction of reactions of each subsystem in each intersection.

We further assessed the metabolic subsystems of the 649 organ-specific metabolic fluxes from the significant associations (**Fig. 3B, Table S2**) and found that most reactions were functionally part of lipid metabolism, consistent with the large number of associations with lipid metabolic features. Reactions of fatty acid metabolism were significantly enriched in associations to blood metabolic features in heart, skeletal muscle, brain and adipose tissue relative to all analysed reactions in each organ-specific metabolic network. In the liver, reactions of glyceride and phospholipid metabolism and vitamin A metabolism were enriched.

There was widespread consistency between biochemical pathways and blood metabolic feature classes (**Fig. 3C, Table S2**). For example, reactions from the glycerides and phospholipids subsystem were primarily associated with blood metabolic features of glycerides and phospholipids as well as lipoprotein fractions and their constituents^20^. We found that reactions of fatty acid metabolism were primarily associated with blood glyceride and phospholipids followed by lipoprotein fractions and fatty acids, which themselves provide acyl chains to glycerides and phospholipids. Similarly, reactions from nucleotide metabolism and amino acid metabolism were primarily associated with blood metabolic features of nucleotides and amino acids, respectively.

### Fluxes of the hepatic triacylglycerol to cholesteryl ester pathway and blood lipoproteins

In the liver, we identified 800 associations between fluxes and lipoprotein fractions (**Table S2**). Many of these associations were to reactions of glycerides and phospholipids metabolism as well as vitamin A metabolism, which were enriched in associations relative to all analysed liver fluxes **(Methods; Fig. 3B)**. FWAS revealed that a major determinant of triacylglycerols (TAG), free cholesterol (FC), and cholesteryl esters (CE) fractions in lipoproteins was a sequential set of reactions which we term the TAG to cholesterol esterification (TAG-CE) pathway (**Fig. 4**). In the TAG-CE pathway, TAGs are hydrolysed to diglycerides and fatty acids in the liver, diacylglycerides are then used as a substrate to synthesise phospholipids (e.g. phosphatidylcholine and phosphatidylethanolamine) which are subsequently used as substrates to esterify FC. We found that fluxes through reactions of the TAG-CE pathway were strongly associated with an increased percentage of CE in HDL and decreased TAG levels in LDL and HDL (**Table 1, Table S3**). The pathway was also strongly associated with reduced HDL size, likely driven by a reduction of TAG levels in HDL^47^. While the associations were primarily found to the liver-specific flux map with mediation by liver-expressed enzymes, these pathways are not necessarily constrained to the liver. For example, the hepatic TAG lipase localises to both liver and blood^48^. Similarly, phospholipids synthesised in the liver may be transferred to HDL in circulation, where they fuel cholesterol esterification catalysed by the liver-secreted lecithin-cholesterol acyltransferase (LCAT)^47,49^.

**Table 1:** Top associations between metabolic fluxes and blood metabolic features in the hepatic triacylglycerol to cholesteryl ester pathway. A complete list of significant associations is provided in **Table S3**. HDL_CE_pct: Cholesteryl esters to total lipids ratio in HDL; HDL_CE_pct_C: Esterified cholesterol to total cholesterol ratio in HDL; HDL_size: Mean diameter for HDL particles; HDL_TG: Triglycerides in HDL; LDL_TG: Triglycerides in LDL.

| *Reaction* | *Blood metabolic feature* | *Effect size* | *FDR-adjusted*<br>*P value*<br>*(INTERVAL)* | *FDR-adjusted*<br>*P value*<br>*(UKB)* |
| --- | --- | --- | --- | --- |
| Triacylglycerol lipase | HDL_CE_pct | + | $2.61 \times 10^{-35}$ | $1.27 \times 10^{-94}$ |
| | HDL_CE_pct_C | + | $1.30 \times 10^{-96}$ | $< 2.225 \times 10^{-308}$ |
| | HDL_size | - | $4.72 \times 10^{-69}$ | $< 2.225 \times 10^{-308}$ |
| | HDL_TG | - | $4.35 \times 10^{-86}$ | $< 2.225 \times 10^{-308}$ |
| | LDL_TG | - | $1.95 \times 10^{-94}$ | $< 2.225 \times 10^{-308}$ |
| Ethanolamine Phosphotransferase | HDL_CE_pct | + | $1.51 \times 10^{-16}$ | $7.20 \times 10^{-36}$ |
| | HDL_CE_pct_C | + | $9.62 \times 10^{-59}$ | $6.53 \times 10^{-232}$ |
| | HDL_size | - | $3.28 \times 10^{-41}$ | $1.38 \times 10^{-236}$ |
| | HDL_TG | - | $2.25 \times 10^{-46}$ | $3.86 \times 10^{-161}$ |
| | LDL_TG | - | $8.08 \times 10^{-55}$ | $3.76 \times 10^{-175}$ |
| Lecithin-Cholesterol Acyltransferase | HDL_CE_pct | + | $1.71 \times 10^{-20}$ | $1.92 \times 10^{-56}$ |
| | HDL_CE_pct_C | + | $7.34 \times 10^{-60}$ | $6.10 \times 10^{-285}$ |
| | HDL_size | - | $2.67 \times 10^{-44}$ | $1.18 \times 10^{-259}$ |
| | HDL_TG | - | $6.78 \times 10^{-53}$ | $1.84 \times 10^{-196}$ |
| | LDL_TG | - | $1.22 \times 10^{-63}$ | $2.27 \times 10^{-211}$ |
| Cholesteryl ester release | HDL_CE_pct | + | $8.62 \times 10^{-29}$ | $2.51 \times 10^{-78}$ |
| | HDL_CE_pct_C | + | $3.88 \times 10^{-81}$ | $< 2.225 \times 10^{-308}$ |
| | HDL_size | - | $3.30 \times 10^{-54}$ | $< 2.225 \times 10^{-308}$ |
| | HDL_TG | - | $4.49 \times 10^{-71}$ | $1.93 \times 10^{-262}$ |
| | LDL_TG | - | $4.85 \times 10^{-80}$ | $2.33 \times 10^{-274}$ |
| Cholesterol Esterase | HDL_CE_pct | - | $1.92 \times 10^{-17}$ | $3.72 \times 10^{-60}$ |
| | HDL_CE_pct_C | - | $2.69 \times 10^{-62}$ | $1.87 \times 10^{-277}$ |
| | HDL_size | + | $6.15 \times 10^{-41}$ | $1.39 \times 10^{-237}$ |
| | HDL_TG | + | $1.05 \times 10^{-47}$ | $8.34 \times 10^{-197}$ |
| | LDL_TG | + | $8.69 \times 10^{-59}$ | $6.68 \times 10^{-208}$ |
| Retinyl Ester Hydrolase | HDL_CE_pct | - | $3.97 \times 10^{-23}$ | $8.50 \times 10^{-71}$ |
| | HDL_CE_pct_C | - | $1.55 \times 10^{-73}$ | $< 2.225 \times 10^{-308}$ |
| | HDL_size | + | $1.02 \times 10^{-48}$ | $1.78 \times 10^{-280}$ |
| | HDL_TG | + | $3.31 \times 10^{-59}$ | $1.97 \times 10^{-233}$ |
| | LDL_TG | + | $8.07 \times 10^{-70}$ | $2.45 \times 10^{-244}$ |

**Fig. 4:**
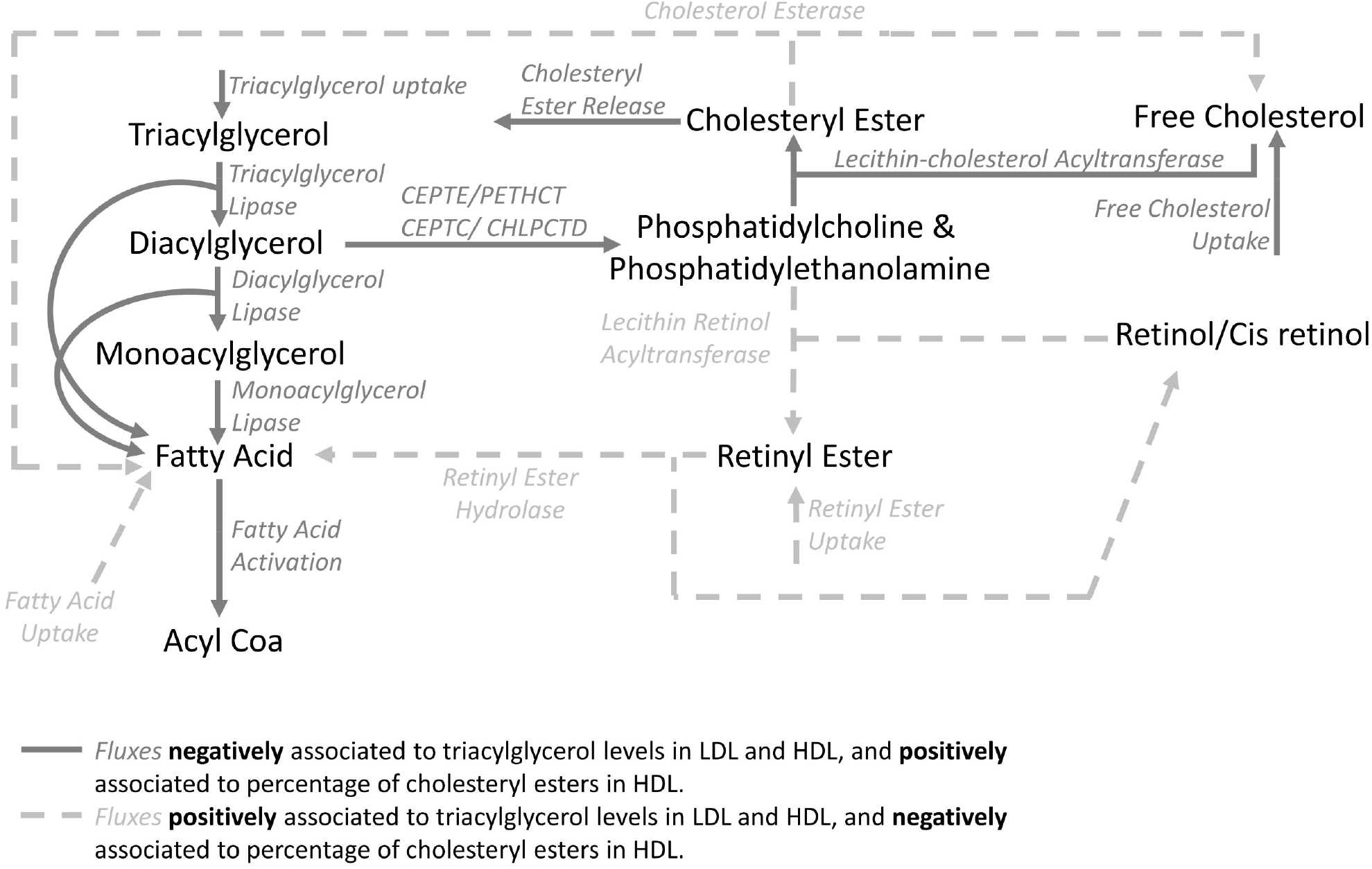
Triacylglycerol to cholesteryl ester pathway in liver. Some transport processes and some metabolites (e.g., glycerol) have been omitted for clarity. CEPTE: Ethanolamine Phosphotransferase; PETHCT: Phosphoethanolamine Cytidyltransferase; CEPTC: Choline Phosphotransferase; CHLPCTD: Choline Phosphate Cytididyltransferase.

Components of the TAG-CE pathway have been the subject of various studies. For example, rare deficiencies in hepatic lipase activity have been linked to increased TAG levels and decreased CE levels in HDL^50^. Similarly, genetic variants in hepatic lipase have been associated with total cholesterol levels in HDL^51,52^. For phospholipids, blocking phosphocholine synthesis has been shown to result in cellar accumulation of TAG both *in vitro* and *in vivo*^53,54^. Similarly, LCAT deficiencies have been associated with reduced cholesterol esterification and increased triglycerides in plasma^55,56^. It has also been suggested that cholesterol is inefficiently esterified by LCAT in patients with CAD, leading to a lower CE to FC ratio^57^.

Conversely, flux through reactions disrupting TAG-CE, such as cholesterol esterase or alternative sources of free fatty acids which lower the flux through triglyceride lipase, are predicted by our FWAS to have the opposite effect and are associated with increased TAG levels and decreased cholesterol esterification (**Table 1, Table S3**). Some of these reactions are related to retinol metabolism (i.e. vitamin A metabolism) as retinols can be esterified by Lecithin Retinol Acyltransferase (LRAT) competing for phospholipids with LCAT, and esterified retinol can act as an alternative source of free fatty acids, thus inhibiting TAG lipase activity **(Fig. 4)**. Notably, administration of high doses of retinol derivatives has been reported to increase total triglyceride levels and, in some instances, increase total cholesterol in LDL while decreasing total cholesterol in HDL^58–61^. We hypothesise that this occurs because retinol derivatives disrupt the hepatic TAG-CE pathway, inhibiting triglyceride lipase and reducing cholesterol esterification, thus reducing the capacity of HDL to collect free cholesterol from other lipoproteins such as LDL^47,57,62^.

### FWAS identifies metabolic fluxes associated with coronary artery disease

We extended our approach of fluxome-wide analysis to common disease and performed a multi-tissue FWAS for coronary artery disease (CAD) in UK Biobank. We associated the 4,418 metabolic fluxes with CAD using a Cox model (**Methods**), which identified 97 significant associations (FDR-adjusted P<0.05 controlling for all tested fluxes). Liver fluxes yielded the largest number of significant associations with CAD (N=46), followed by fluxes of adipose tissue (21), brain (12), heart (10) and skeletal muscle (8) (**Fig. 5**; **Table S4**).

**Fig. 5:**
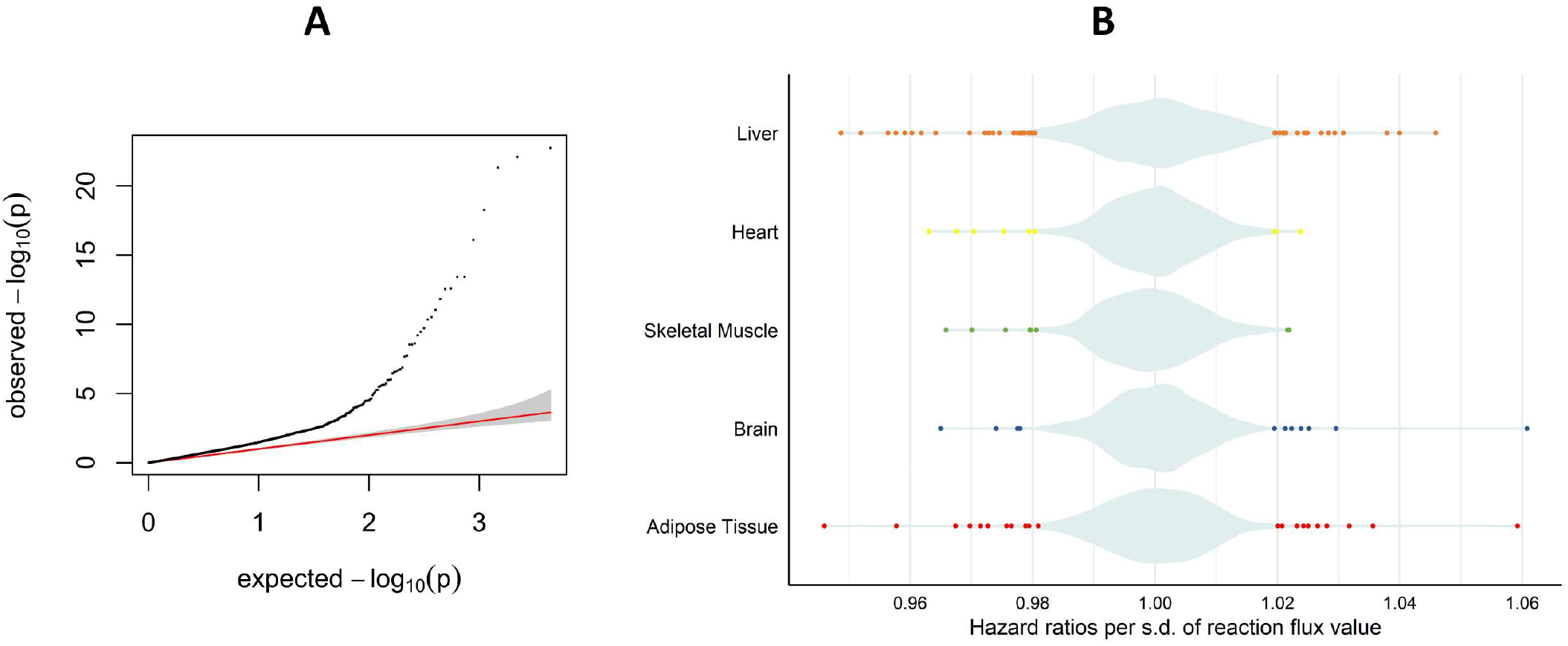
Fluxome-wide association analysis (FWAS) between genetically personalised flux maps and coronary artery disease. (**A**) quantile-quantile (QQ) plot of the observed P values for associations between flux values and CAD risk. (**B**) Plot of the significant reaction flux hazard ratios per organ on coronary artery disease risk. The violin plot, coloured in pale azure, shows the distribution of both significant and non-significant hazard ratios.

Notably, the strongest association with CAD was the flux of the transport of norepinephrine in the brain with a hazard ratio (HR) per s.d. of log-transformed flux value of 1.060 and a P value of 1.9×10^−23^ (**Table S4**). Norepinephrine is a neurotransmitter that can increase blood pressure and may play a role in atherosclerosis^63,64^. Fluxes through reactions of histamine metabolism were shown to have a strong association with CAD in adipose tissue (release of 1-methylhistamine: HR=1.059 per s.d., P= 8.30×10^−23^) and liver (histamine release: HR=1.046 s.d., P=3.60×10^−14^, histidine uptake: HR=1.040 per s.d., P= 3.00×10^−11^); Histamine is an inflammatory mediator synthesised from histidine primarily in mast cells^65^, which reside in most tissues, including liver and adipose tissue^66,67^. Histamine has been reported to be associated with atherosclerosis via blood lipids and lipoprotein fractions as well as by promoting inflammation^68–71^. Conversely, flux through the triacylglycerol lipase reactions in adipose tissue (HR=0.95771 per s.d., P= 2.90×10^−13^), heart (HR= 0.963 per s.d., P= 1.90 ×10^−10^), skeletal muscle (HR= 0.966 per s.d., P= 2.90×10^−9^) were strongly associated with reduced CAD risk, consistent with the anti-atherogenic effect of lipoprotein lipase activity in these organs^72^.

In the liver, fluxes through 29 transport processes were significantly associated with CAD. Fifteen of which were mediated by the transmembrane receptor SLCO1B1, including the transport of arachidonic acid (HR per s.d. = 0.949, P=5.4×10^−19^), prostaglandins (e.g. transport of prostaglandin I2: HR per s.d.= 0.952, P= 8.10 ×10^−17^), and bile acids (e.g. transport of glycocholic acid: HR per s.d.= 0.958, P=5.40×10^−19^). SLCO1B1 mediates the uptake of both endogenous and xenobiotic compounds into hepatocytes. Notably, SLCO1B1 also mediates the hepatic uptake of statins, enhancing their therapeutic efficacy^73,74^. The flux of arachidonic acid secretion to the bile was also associated with reduced CAD risk (HR per s.d.=0.974, P= 5.40×10^−6^). Arachidonic acid is a precursor of many prostaglandins and has been associated with the risk of atherosclerosis, most recently through Mendelian randomisation^75,76^. Similarly, SLCO1B1-independent transport of bile acids was also associated to decreased CAD risk, such as flux through the antiport of deoxycholic acid and carbonate (HR per s.d.= 0.973, P=3.20×10^−6^) and the ATP-binding cassette mediated efflux of tauroursodeoxycholate (HR per s.d.= 0.977, P= 8.00×10^−5^). Bile salt excretion enables the liver to remove excess cholesterol and is reported to be reduced in CAD patients compared to healthy controls^77^.

In adipose tissue, seven reactions of fatty acid metabolism were associated with CAD risk, such as the transport of docosahexaenoic acid from mitochondria to the cytosol (HR per s.d.=0.94595, P=4.80×10^−22^), the activation of myristic acid (HR per s.d.= 0.968, P= 1.90×10^−8^) and its transport to the mitochondria as tetradecanoyl carnitine (HR per s.d.=0.972, P=1.10×10^−6^). Notably, saturated fatty acids, like myristic acid, have been associated with CAD risk^78,79^, whereas polyunsaturated fatty acids such as docosahexaenoic are negatively correlated with CAD risk and severity^80,81^. This suggests that fatty acid metabolism in adipose tissue might influence CAD risk by modulating the bioavailability of pro- and anti-atherogenic fatty acids. Further, in adipose tissue, the flux through the phospholipase reaction that hydrolyses phospholipids was associated with increased CAD risk (HR per s.d.= 1.032, P=1.30×10^−7^). Phospholipase activities have been suggested to have a causal role in atherosclerosis and have been investigated as potential pharmacological targets to prevent atherosclerosis and CAD^82–85^.

## Discussion

Here we present a new framework that uses metabolic modelling to leverage the stoichiometric relationships of enzymes in human genome-scale metabolic networks to characterise how genetic variants affect metabolic phenotypes. We achieve this by integrating genetic effects on transcript levels into organ-specific GSMMs and simulating how they propagate and interact into genome-scale flux maps of major human organs. To validate our method, we built organ-specific models for adipose tissue, brain, heart, liver and skeletal muscle for over 520,000 individuals from the INTERVAL^34,35^ and UKB^36^ cohorts, increasing by more than two orders of magnitude the number of personalised GSMMs built in previous works^29–32^. Association analyses were performed between genetically-personalised flux values and directly measured blood metabolites in both INTERVAL and UKB, identifying many significant and replicable associations. As expected, we found that most blood metabolic features were associated with functionally related flux pathways. Finally, we demonstrate that fluxome-wide analysis can be used to identify putative metabolic drivers of coronary artery disease.

With cardiovascular disease a leading cause of mortality and comorbidity worldwide, the identification of specific biochemical reactions using population-scale genomic data is of significant interest to both basic discovery science and the development of therapeutics^86^. Many of the 97 flux associations we identified involve pathways or metabolites that have been associated with CAD in existing studies, such as histamine metabolism^68,71^, arachidonic acid^75,76^, docosahexaenoic acid^80,81^, triacylglycerols^87^ or the phospholipase activity^82,83,85^. Modulation of some of these fluxes have been explored as therapies for CAD, namely several phospholipase inhibitors^83,84^.

As a proof of concept, this study focused on modelling five of the most prominent human organs^39^. Given the increasing availability of models to impute tissue- or cell-specific transcript abundance from genotype^12^, this approach may be expanded to other tissues and cell types. We envision that future applications may also select organs for modelling based on the target disease(s). Similarly, the modelling framework could also be expanded to include additional layers of data such as imputed protein abundance^14^ and gain or loss of function mutations^88^.

GSMMs have previously been shown to have utility for drug discovery and repositioning^31,89–91^. Therefore, FWAS, based on genetically-personalised organ-specific models, may identify fluxes associated with disease states and, by extension, the gene knockdowns or metabolic interventions (e.g. dietary supplements or metabolic inhibitors) to target them. FWAS to blood metabolic features may also help screen potentially adverse side effects of metabolic interventions. For example, we identified that retinols may increase TAG levels and reduce cholesterol esterification in lipoproteins, consistent with reports that administering high doses of vitamin A derivatives results in hypertriglyceridemia and dysregulation of cholesterol levels^58–61^.

Overall, this work demonstrates that genome-scale metabolic modelling can contribute to addressing the V2F challenge by characterising how the effects of genetic variants propagate through metabolic networks of specific human organs.

## Methods

### INTERVAL

INTERVAL is a cohort of approximately 50,000 participants nested within a randomised trial studying the safety of varying the frequency of blood donation. Participants were blood donors aged 18 years and older (median 44 years of age; 50% women) recruited between 2012 and 2014 from 25 NHS Blood and Transplant centers^34,35^. Genotyping of INTERVAL samples and their quality control and imputation were performed as previously described^92^.

### UK Biobank

UKB is a cohort of approximately 500,000 participants from the general UK population. Participants were between age 40 and 69 years at recruitment (median 58 years of age; 54% women) and accepted an invitation to attend one of the assessment centres that were established across the United Kingdom between 2006 and 2010^36^. Genotyping, quality control and imputation of UKB samples were performed as previously described^92^.

### Metabolomics

The Nightingale NMR platform quantifies 230 and 249 analytes in INTERVAL and UKB, respectively, including lipoprotein subfractions and ratios, lipids and low molecular weight metabolites (e.g., amino acids). Values were adjusted for technical covariates as previously described^93^ and subsequently regressed for age, sex, BMI, and the first 5 PCs of genetic ancestry, with composite biomarkers and ratios recomputed after adjustment, including 98 and 76 additional biomarker ratios in INTERVAL and UKB, respectively, not provided by the Nightingale platform^93^. Eight measures present in INTERVAL but not UKB were excluded. Likewise, 68 features with markedly distinct variance between INTERVAL and UKB (|log2(sd_INTERVAL_/sd_UKB_)|> log2(2.5)) were also excluded. Finally, acetate was excluded due to a large number of NA(>75%) in INTERVAL. Subsequently, measures were standardised to zero-mean and unit variance.

The Metabolon HD4 assay measures ∼1000 metabolites (∼700 named, ∼300 unknown), including lipids, xenobiotics, amino acids and energy-related metabolites. Nineteen features were excluded due to a large number of NA(>75%). Values were regressed against technical covariates age, sex, BMI, and the first 5 PCs of genetic ancestry. Subsequently, measures were standardised to zero-mean and unit-variance.

### Building organ-specific reference flux maps

For each analysed organ (i.e., adipose tissue, brain, liver, heart and skeletal muscle), the organ-specific genome-scale metabolic network was extracted from the Harvey/Harvetta metabolic reconstructions^29^. To avoid any gender biases, any reaction present in either the male (Harvey) or female (Harvetta) models was included, and the reaction flux boundaries for exchange reactions were established as the average between both models. Metabolites in blood or bile ducts were made boundary conditions (i.e., assumed constant), allowing each organ subnetwork to function independently. Lastly, networks were manually tuned to ensure that the resulting flux maps were biologically relevant and that each organ could adequately fulfil its basic metabolic functions. The process involved modifying the constraints of exchange reactions so that the ranges of metabolite uptake and secretion of each organ were physiologically feasible, filling gaps in the pathways of fatty acid metabolism and removing artificial sink reactions. Additionally, the artificial metabolites Rtotal, used to simulate the side acyl chains of triacylglycerides in the RECON family of GSMMs^25^, were substituted by a stoichiometric mix of 1/3 oleic acid, 1/6 palmitoleic acid, 1/6 palmitic acid, 1/6 stearic acid and 1/6 myristic acid. This was necessary to ensure that mass was conserved while allowing both *novo*-synthesis and degradation of triacylglycerols. The resulting metabolic networks are available at https://github.com/cfoguet/cobrafunctions/tree/main/example_inputs/organ_specific_models.

Next, a set of metabolic requirements were defined for each organ representing metabolic functions that each organ fulfils in physiological conditions (**Table S5**). These were added in each organ subnetwork as lower bounds for flux values through reactions associated with those metabolic requirements. Lower bounds were set relative to the maximum flux feasible through such reactions identified with flux variability analysis(FVA)^94^. In addition, all organs were also constrained to be able to maintain their biomass (i.e., to be able to synthesise structural lipids, proteins, and nucleotides to counter their natural decay).

Organ-specific transcript abundances were obtained as Transcripts Per Million (TPM) from the GTEx Portal^13^ on 05/05/2021 (GTEx Analysis Release V8; dbGaP Accession phs000424.v8.p2) and the average abundance of each transcript in each organ was obtained. In the heart, adipose tissue, and brain, there were transcripts abundances measured from multiple source sites. Clustering analysis indicated that samples from each organ source site were clustered together (**Fig. S7**). Hence, the average of all sample source sites was used for heart, adipose tissue and brain tissue.

Average transcript abundances were mapped to the organ-specific subnetworks using the gene reaction annotations of Harvey/Harvetta^29^. More in detail, transcript abundances of isoenzymes and enzymes subunits catalysing each reaction or transport process were added and, subsequently, log2 transformed. The resulting values were used as input to apply the GIM3E algorithm^37^. The GIM3E algorithm applies a flux minimisation weighted by transcript abundance allowing to identify solutions that are both enzymatically efficient and consistent with gene expression data (**Fig. S1**).

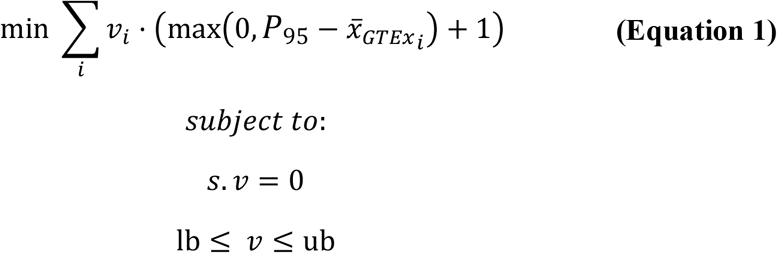

where,

*ν* is a vector of steady-state flux values; 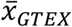 is a vector of average transcript abundances mapped to reactions of the organ-specific network; *P*_95_ is the 95^th^ percentile of the average transcript abundance values mapped to reactions of the organ-specific network; *s* is the stoichiometric matrix. Its product with *ν* defines the metabolic steady state constraint (i.e., input and output fluxes must be balanced for each metabolite in the network); *lb* and *ub* are vectors defining the lower and upper bounds of reactions, respectively. The organ-specific metabolic requirements are defined as lower bounds greater than 0 (i.e., constraining such reactions to being active).

Subsequently, FVA was used to identify the feasible flux ranges within 99% of the optimal value of the GIM3E objective function ^37^. Finally, the resulting solution space was sampled using the Artificially Centered hit-and-run (ACHR) algorithm implemented into COBRApy^95,96^. ACHR was run with a thinning factor of 1000, and 1000 sets of steady-state flux distributions were computed. The average of those flux samples was used as each organ’s reference flux map.

### Imputing individual-specific gene expression data

The GTEx v8 elastic net models from PredictDB (https://predictdb.org) were used to impute organ-specific gene expression levels from individual-level genotypes. The models were used with PLINK2^97^ to predict relative transcripts abundances using genotype data from INTERVAL^34,35^ and UK Biobank^36^ cohorts. For adipose, brain and heart tissue, the average of the imputed abundances in each source site were used.

### Mapping individual-specific gene expression data to reactions in the model

Imputed individual-specific expression patterns from metabolic genes (i.e., genes coding for enzymes, enzyme subunits, or transmembrane carriers) were mapped to organ-specific models using the gene reaction annotations of Harvey/Harvetta^29^. Imputed values were expressed as log_2_ fold changes relative to average gene expression in GTEx. Imputed fold changes were then mapped to reactions in the organ-specific model considering the relative transcript abundance of isoenzymes and enzyme subunits in GTEx:

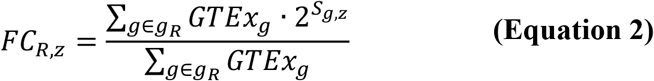

where,

*S*_*g,z*_ is the organ-specific score for gene *g* in individual *z* computed using the elastic nets models from PredictDB; *GTEx*_*g*_ is the average organ-specific gene expression in GTEx of gene *g; g*_*R*_ are the genes associated with reaction *R* in the organ-specific network; *FC*_*R,z*_ is the imputed reaction activity fold change for reaction *R* in individual *z*.

### Quadratic metabolic transformation algorithm (qMTA)

Building upon the principle of the metabolic transformation algorithm^91,98^, we developed qMTA. qMTA seeks to identify the flux map most consistent with a set of reaction activity fold changes starting from a reference flux distribution (**Equation 3**; **Fig. S2**). To this end, it minimizes the difference between the simulated flux values and the target fluxes (i.e. the product of the flux value in the reference flux distribution and the reaction activity fold change). Additionally, it also minimizes the deviation from the reference flux distribution in reactions not mapped to any gene expression fold changes. Furthermore, the two terms of the optimization function are scaled by the reference flux distribution to prevent biases towards reactions with high flux values.

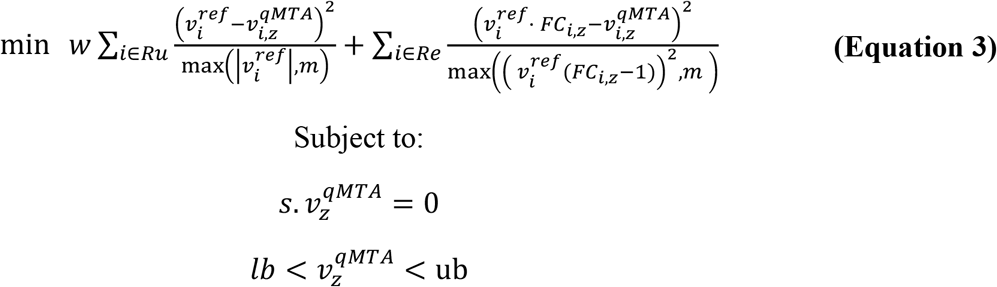

where,

*w* is the weight given to minimising variation in reactions not mapped to imputed gene expression;*Ru* are reactions not mapped to imputed gene expression;*ν*^*ref*^is the flux vector of the reference flux distribution; 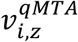 is the simulated flux value for reaction *i* in individual *z*; *m* is the minimum value allowed for the scaling factor;*Rg* are reactions mapped to imputed gene expression.

Personalised flux maps computed with 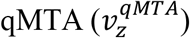 were subsequently log_2_ transformed and standardized to zero-mean and unit variance.

Additionally, two hyperparameters in qMTA (*w* and *m*) were tuned using the regression analysis with blood metabolic features in the INTERVAL cohort. For each simulated organ, a grid search (*w*→[100,10,1,0.1,0.01], *m*→ [10^−6^, 10^−7^, 10^−8^, 10^−9^, 10^−10^, 10^−11^, 10^−12^]) was performed to identify the parameters that resulted in flux maps with the strongest association with both Nightingale Health and Metabolon HD4 metabolic features. This was measured as the summation of the amount of variance explained (R^2^) for each pair of blood metabolic feature and flux value when testing associations between metabolic fluxes and blood metabolic features. The resulting parameters were subsequently used in the analysis of the samples from UKB.

### Testing associations between metabolic fluxes and blood metabolic features

Due to the linear nature of many metabolic pathways, some flux values were highly intercorrelated (**Fig S3-S4)**. To remove reaction flux pairs with a strong correlation, for each pair of reaction flux values with ρ>0.9, the feature with the largest mean absolute correlation to other flux values was removed^99^. Likewise, both the Nightingale and Metabolon platforms had some metabolic features with strong correlations and those features with ρ>0.75 were removed using the same approach used for reaction fluxes. Overall, 4,418 scaled flux values and 57 Nightingale Health and 713 Metabolon HD4 blood metabolic features were selected to perform FWAS.

Then, the association of each metabolic feature to each personalised flux value was evaluated using a linear regression (**Equation 4**;**Fig. S2**).

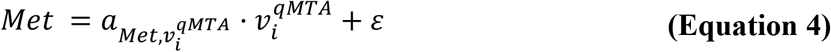

where,

*Met* are the measured levels of a blood metabolic feature;

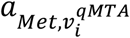 is the effect size of flux *i* on *Met*;

*ε* is the residual.

Statistical significance was evaluated with a t-test on effect sizes. The resulting p-values were adjusted for multiple testing against all evaluated blood metabolic features – reaction flux pairs using the Benjamini and Yosef Hochberg (i.e., FDR) method.

### Classes of blood metabolic features

Nightingale/Metabolon platforms provide sets of Groups/Sub pathways to stratify metabolic features. We harmonised both annotations systems to define a set of curated groups that could be applied to both Nightingale and Metabolon features (**Table S6**). For instance, the Metabolon features annotated to “Glycerolipid Metabolism” and “phospholipid metabolism”, and the Nightingale features annotated to “phospholipids” were all reassigned to the curated group “Glycerides and phospholipids”. Some Metabolon features were not annotated (i.e., unknown) and could not be assigned to any curated group. Unknown features were included in the FWAS but omitted from enrichment analysis. Fisher’s exact test was used to identify metabolite classes enriched in features with significant association to personalised flux values relative to the set of all uncorrelated blood metabolic features. An FDR-adjusted significance threshold of P<0.05 was applied to control for all tested classes of blood metabolic features across all organs.

### Reaction subsystems

Subsystem annotations for reactions were obtained from the Harvey/Harvetta model^29^. As some subsystems either contained an excessively low number of reactions or had reactions that functionally could be considered to belong to several subsystems, similar subsystems were aggregated into large ones. For instance, pyrimidine synthesis, purine synthesis, pyrimidine catabolism, purine catabolism and nucleotide interconversion were aggregated into a subsystem termed nucleotide metabolism. Additionally, reactions from transport subsystems were reassigned to the reaction subsystem most frequent in the metabolites being transported (e.g., transport of palmitate across the extracellular membrane was reassigned from “Transport, extracellular” to “Fatty acid metabolism”) (**Table S7**). Fisher’s exact test was used to identify subsystems enriched in reactions with significant association with blood metabolic features relative to the set of all evaluated reactions in a given organ. An FDR-adjusted significance threshold of P<0.05 was applied to control for all tested subsystems across all organs.

### Cox’s proportional hazard model

The association of genetically personalised fluxes to CAD was evaluated in individuals of British ancestry in UKB. Using PheWAS Catalog (version 1.2), we used the WHO International Classification of Diseases (ICD) diagnosis codes in versions 9 (ICD-9) and 10 (ICD-10) of Phecode 411.1 for CAD case definition in UKB. In detail, we searched for the presence of any of the constituent ICD-9/10 codes in linked health records (including in-patient Hospital Episode Statistics data, and primary and secondary cause of death information from the death registry), and converted the earliest coded date to the age of phenotype onset. Individuals without any codes for the phenotype of interest were recorded as controls and censored according to the maximum follow-up of the health linkage data (January 31, 2020) or the date of death. Thus, to define the cohort for testing flux associations with the age-of-onset of CAD, we used the set of events and censored individuals described above.

Association was evaluated using the age-as-time-scale Cox proportional hazards regression. The Cox models were stratified by sex and adjusted by genotyping array, 10 genetic PCs, BMI and smoking status. The Cox model was fitted using the CoxPHFitter function from the lifelines package for python^100^.

## Data Availability

All data produced in the present study are available upon reasonable request to the authors

## Code Availability

The workflow to build personalised organ-specific flux maps from imputed gene expression data uses custom-built functions and scripts implemented in the framework of COBRA for python(COBRApy)^95,96^. qMTA requires the proprietary solver CPLEX, which is freely available to academic users. Code is available at https://github.com/cfoguet/cobrafunctions.

## Acknowledgements

Participants in the INTERVAL trial were recruited with the active collaboration of NHS Blood and Transplant (www.nhsbt.nhs.uk), which has supported fieldwork and other elements of the trial. DNA extraction and genotyping were co-funded by the National Institute for Health Research (NIHR), the NIHR BioResource (http://bioresource.nihr.ac.uk) and the NIHR Cambridge Biomedical Research Centre (BRC) (no. BRC-1215-20014)*. Nightingale Health NMR assays were funded by the European Commission Framework Programme 7 (HEALTH-F2-2012-279233). Metabolon Metabolomics assays were funded by the NIHR BioResource and the NIHR Cambridge Biomedical Research Centre (BRC-1215-20014)*.The academic coordinating centre for INTERVAL was supported by core funding from the NIHR Blood and Transplant Research Unit in Donor Health and Genomics (no. NIHR BTRU-2014-10024), UK Medical Research Council (MRC) (no. MR/L003120/1), British Heart Foundation (nos SP/09/002, RG/13/13/30194 and RG/18/13/33946) and the NIHR Cambridge BRC (no. BRC-1215-20014)*. *The views expressed are those of the author(s) and not necessarily those of the NIHR or the Department of Health and Social Care. A complete list of the investigators and contributors to the INTERVAL trial is provided in ref. ^34^. The academic coordinating centre would like to thank blood donor centre staff and blood donors for participating in the INTERVAL trial. This work was supported by Health Data Research UK, which is funded by the UK MRC, Engineering and Physical Sciences Research Council (EPSRC), Economic and Social Research Council, Department of Health and Social Care (England), Chief Scientist Office of the Scottish Government Health and Social Care Directorates, Health and Social Care Research and Development Division (Welsh Government), Public Health Agency (Northern Ireland), British Heart Foundation and Wellcome. The authors are grateful to UK Biobank for access to data to undertake this study (Projects #7439). This work was performed using resources provided by the Cambridge Service for Data Driven Discovery (CSD3) operated by the University of Cambridge Research Computing Service (www.csd3.cam.ac.uk), provided by Dell EMC and Intel using Tier-2 funding from the Engineering and Physical Sciences Research Council (capital grant EP/P020259/1), and DiRAC funding from the Science and Technology Facilities Council (www.dirac.ac.uk).

## Supplementary Figures and Tables

**Fig. S1: Computing the organ-specific reference flux distribution**. First, the space of solutions consistent with reaction stoichiometry from the organ subnetwork is extracted from the Harvey/Harvetta multiorgan model. *s* is the stoichiometric matrix, *ν* is a vector of steady-state flux values and *lb* and *ub* the vector of reaction lower and upper bounds, respectively. An example of the stoichiometric matrix for a reaction network with three reactions and one ramification is provided, and a hypothetical feasible space is shown on a 3D space. Next, a set of organ-specific metabolic functions are defined, and the solution space is constrained to only solutions above a given threshold (*lb*_*obj*_) for these objectives. Next, a vector of minimisation weights (*w*) for reactions is defined using average transcript abundances from GTEx mapped to reactions of the organ-specific network 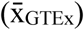. P_95_ is the 95^th^ percentile of the average transcript abundance values mapped to reactions of the organ-specific network. Next, the GIM3E algorithm is used to perform a weighted minimisation of total reaction fluxes. Finally, the vicinity of the GIM3E solution space is defined and sampled to obtain a representative flux map. *Th*_GIM3E_ is the maximum value of the GIM3E objective used to define the vicinity of the GIM3E solution space.

**Fig. S2: Genetically personalised flux values and fluxome-wide association studies (FWAS)**. First, personalised organ-specific transcript abundances are imputed from genotype data and mapped to reactions in the organ-specific metabolic network as reaction activity fold changes. Next, qMTA is used to find the flux distribution most consistent with the reaction activity fold changes in each individual, starting from the reference flux distribution. *v*^ref^ is the flux vector of the reference (average) flux distribution. *Re* are reactions mapped to imputed gene expression. *Ru* are reactions not mapped to imputed gene expression. FC_R,z_ is the reaction activity fold change imputed from genotype data for reaction R in in individual *z*. For clarity, some parameters have been omitted from the qMTA equation, the full equation can be found in Methods. The resulting personalised flux values can be used to perform FWAS to complex traits or diseases. As part of this process, a given trait is independently regressed against the fluxes through each reaction in the network to test for association. For instance, to perform FWAS on blood metabolome, the measure of each metabolite will be linearly regressed against the flux through each reaction in a given organ metabolic network. Blood metabolic features can include the concentration of metabolites (e.g., [Glc]: Glucose concentration), the concentration of metabolite fractions in lipoproteins (e.g. [LDL_TG]: Triglycerides in LDL) and ratios and relative measures (e.g., *Unsat*%: degree of fatty acid unsaturation).

**Fig. S3: Example of correlation between fluxes in a set of reactions in the liver**. (A) Set of interconnected reactions centred around triacylglycerol. (B) Density plots for flux values through the reactions in the pathway. (C) Heatmaps of pairwise Pearson correlation coefficients between reaction flux values in the pathway. Stoichiometric coupling between reactions in the pathway (i.e., reactions sharing substrates or products) results in a correlation between flux values through the reactions. CEPTC: Choline Phosphotransferase; CEPTE: Ethanolamine Phosphotransferase; FA uptake: Fatty acid uptake; FACOAL: Fatty Acid CoA synthase. LPS: Triacylglycerol lipase; LPS2: Diacylglycerol lipase; LPS3: Monoacylglycerol lipase; RETH: Retinyl Ester Hydrolase.

**Fig. S4: Heatmaps of pairwise Pearson correlation coefficients between reaction flux values before and after filtering out reactions with strong pairwise correlations**. Reaction fluxes in each organ are ordered based on hierarchical clustering of correlation coefficients. To remove reaction flux pairs with strong correlation, for each pair of reaction flux values with ρ>0.9 the feature with the largest mean absolute correlation to other flux values was removed.

**Fig. S5: Principal component analysis**. Pairwise score plots between the first five principal components in UKB and INTERVAL computed using genetically personalised metabolic flux values as features. Each organ is analysed and plotted independently.

**Fig. S6: Plot of flux effect sizes per organ to blood metabolic features measured with the Nightingale Health platform in INTERVAL and UKB**.

**Fig. S7: Clustering of GTEx source sites**. Source **s**ites are clustered hierarchically using the correlation between average transcripts abundance in each site as a measure of similarity.

**Table S1: Principal component loadings of genetically personalised flux values across all modelled organs**. The ten flux values with the most contribution to the first five principal components are indicated. Each organ is analysed independently.

**Table S2: FWAS to blood metabolic features**. A table detailing the 4,411 significant associations identified in the INTERVAL cohort between genetically personalised flux values and blood metabolic features.

**Table S3: Extended FWAS to lipoprotein fractions in the liver**. Once FWAS analysis had established the importance of liver fluxes in lipoprotein fractions, FWAS was repeated using all fluxes from the liver (i.e. without excluding fluxes with strong intercorrelation), and all lipoproteins blood features (i.e. without excluding features with strong intercorrelation) to provide a complete picture. The table details the 16,356 significant associations in the INTERVAL cohort between all genetically personalised flux values in the liver and all lipoprotein features.

**Table S4: FWAS to coronary artery disease**. Table detailing the 97 genetically personalised flux values with significant associations to coronary artery disease risk in the UK Biobank cohort.

**Table S5: Metabolic requirements of each organ**.

**Table S6**: **Annotations for blood metabolic features in the Nightingale and Metabolon platforms**. Nightingale/Metabolon platforms each provide sets of Groups/Sub pathways to stratify metabolic features. Both annotations systems were harmonised to define a set of curated groups that could be applied to both Nightingale and Metabolon features.

**Table S7**: **Metabolic subsystem annotations for reactions in the organ-specific genome-scale metabolic models**.

