## Supplemental Figures for "Genetically personalised organ-specific metabolic models in health and disease"

### Supplementary Figures

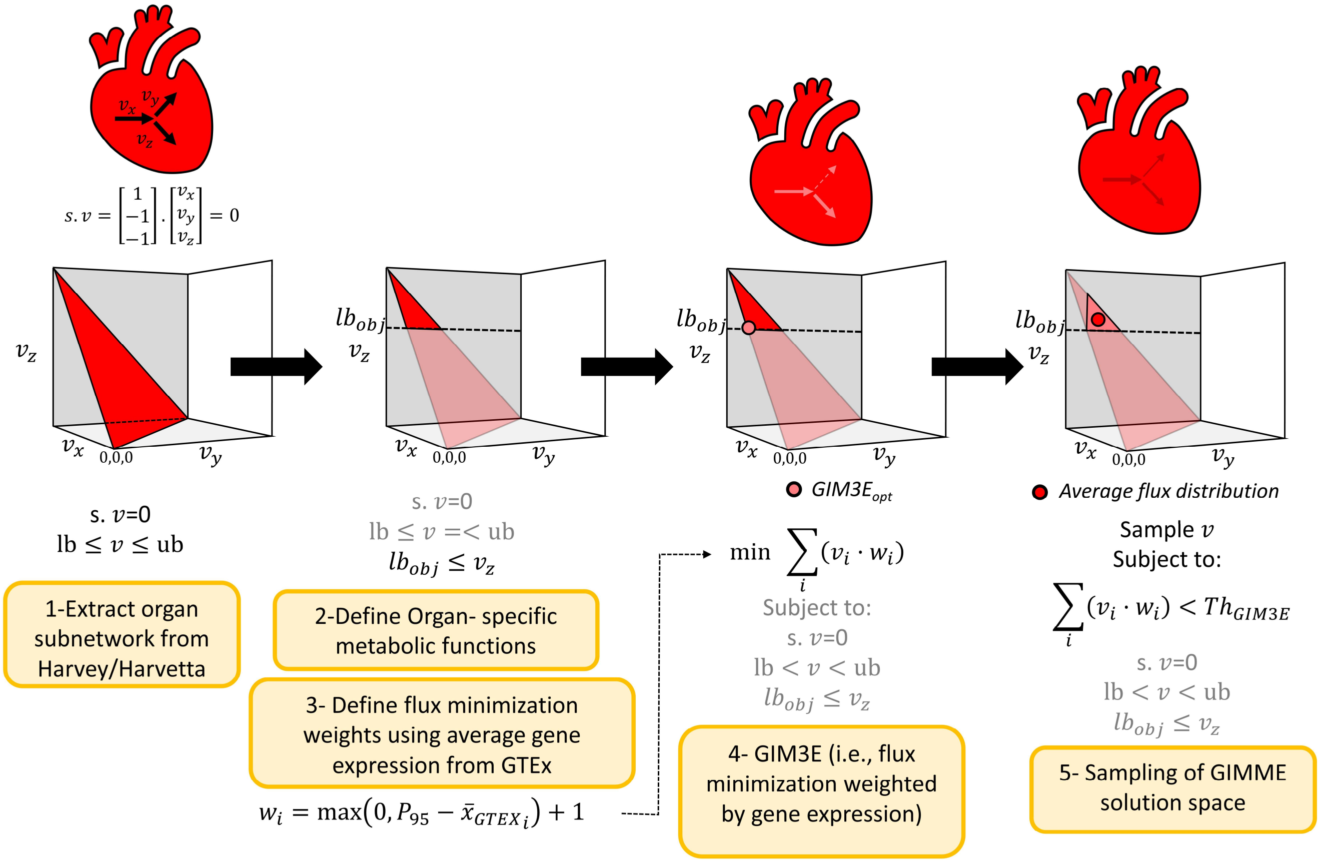


**Fig. S1: Computing the organ-specific reference flux distribution**. First, the space of solutions consistent with reaction stoichiometry from the organ subnetwork is extracted from the Harvey/Harvetta multiorgan model. *s* is the stoichiometric matrix, *v* is a vector of steady-state flux values and *lb* and *ub* the vector of reaction lower and upper bounds, respectively. An example of the stoichiometric matrix for a reaction network with three reactions and one ramification is provided, and a hypothetical feasible space is shown on a 3D space. Next, a set of organ-specific metabolic functions are defined, and the solution space is constrained to only solutions above a given threshold ($lb_{obj}$) for these objectives. Next, a vector of minimisation weights ($w$) for reactions is defined using average transcript abundances from GTEx mapped to reactions of the organ-specific network ($\bar{x}_{GTEx}$). $P_{95}$ is the 95$^{th}$ percentile of the average transcript abundance values mapped to reactions of the organ-specific network. Next, the GIM3E algorithm is used to perform a weighted minimisation of total reaction fluxes. Finally, the vicinity of the GIM3E solution space is defined and sampled to obtain a representative flux map. $Th_{GIM3E}$ is the maximum value of the GIM3E objective used to define the vicinity of the GIM3E solution space.

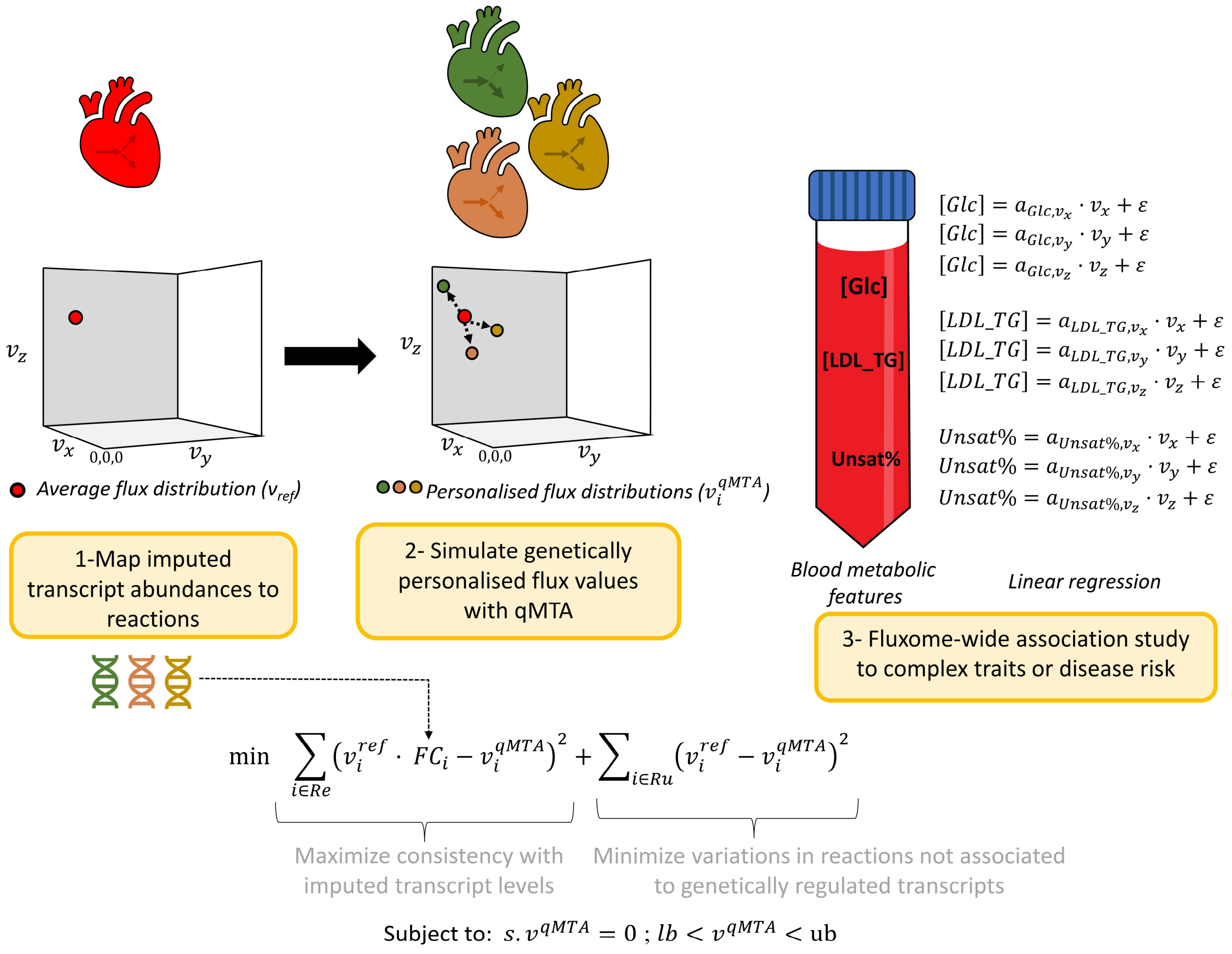


**Fig. S2: Genetically personalised flux values and fluxome-wide association studies (FWAS)**. First, personalised organ-specific transcript abundances are imputed from genotype data and mapped to reactions in the organ-specific metabolic network as reaction activity fold changes. Next, qMTA is used to find the flux distribution most consistent with the reaction activity fold changes in each individual, starting from the reference flux distribution. $v^{ref}$ is the flux vector of the reference (average) flux distribution. *Re* are reactions mapped to imputed gene expression. *Ru* are reactions not mapped to imputed gene expression. $FC_{R,z}$ is the reaction activity fold change imputed from genotype data for reaction R in in individual *z.* For clarity, some parameters have been omitted from the qMTA equation, the full equation can be found in **Methods.** The resulting personalised flux values can be used to perform FWAS to complex traits or diseases. As part of this process, a given trait is independently regressed against the fluxes through each reaction in the network to test for association. For instance, to perform FWAS on blood metabolome, the measure of each metabolite will be linearly regressed against the flux through each reaction in a given organ metabolic network. Blood metabolic features can include the concentration of metabolites (e.g., [Glc]: Glucose concentration), the concentration of metabolite fractions in lipoproteins (e.g. [LDL_TG]: Triglycerides in LDL) and ratios and relative measures (e.g., *Unsat%*: degree of fatty acid unsaturation).

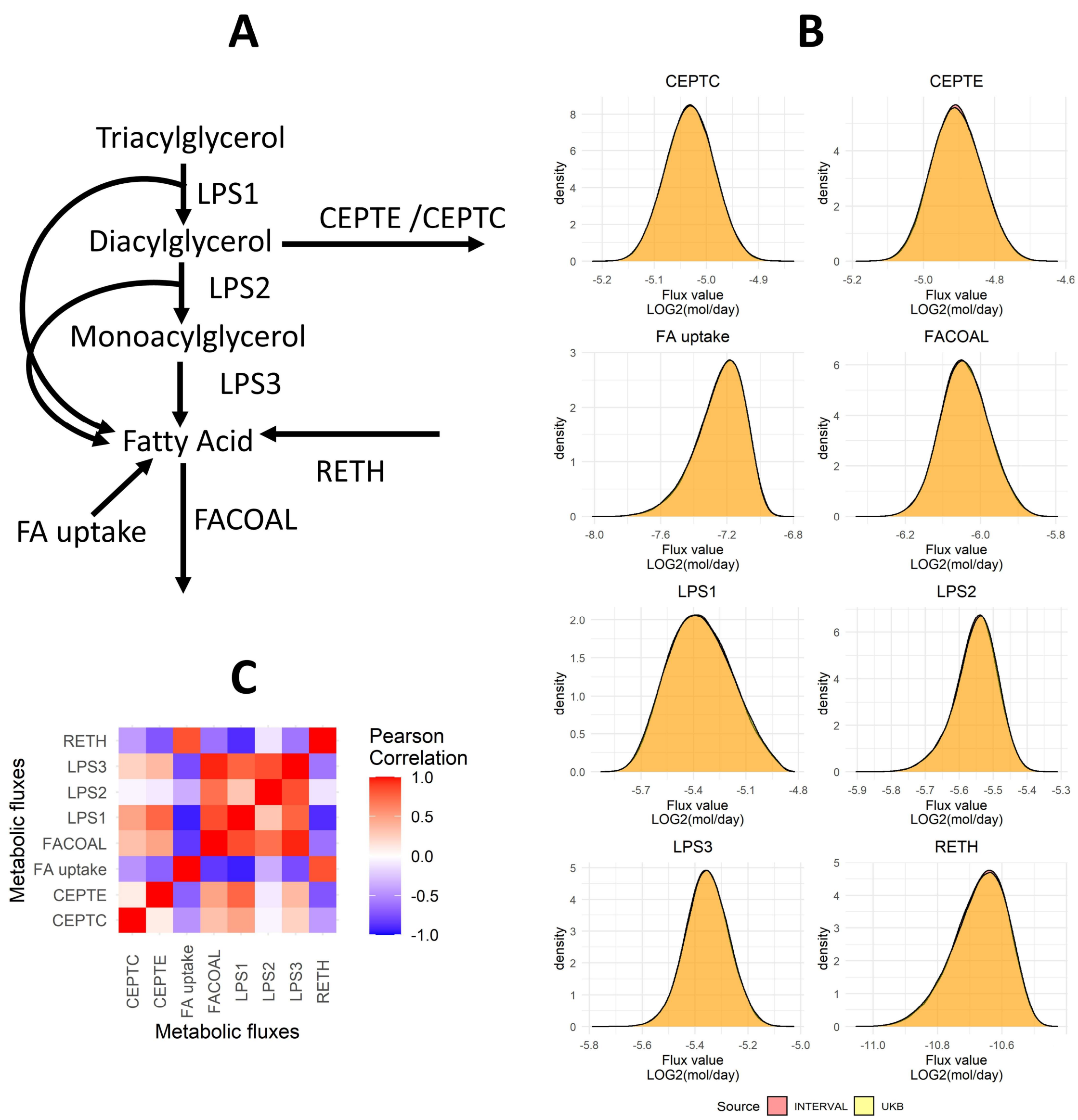


**Fig. S3: Example of correlation between fluxes in a set of reactions in the liver**. (A) Set of interconnected reactions centred around triacylglycerol. (B) Density plots for flux values through the reactions in the pathway. (C) Heatmaps of pairwise Pearson correlation coefficients between reaction flux values in the pathway. Stoichiometric coupling between reactions in the pathway (i.e., reactions sharing substrates or products) results in a correlation between flux values through the reactions. CEPTC: Choline Phosphotransferase; CEPTE: Ethanolamine Phosphotransferase; FA uptake: Fatty acid uptake; FACOAL: Fatty Acid CoA synthase. LPS: Triacylglycerol lipase; LPS2: Diacylglycerol lipase; LPS3: Monoacylglycerol lipase; RETH: Retinyl Ester Hydrolase.

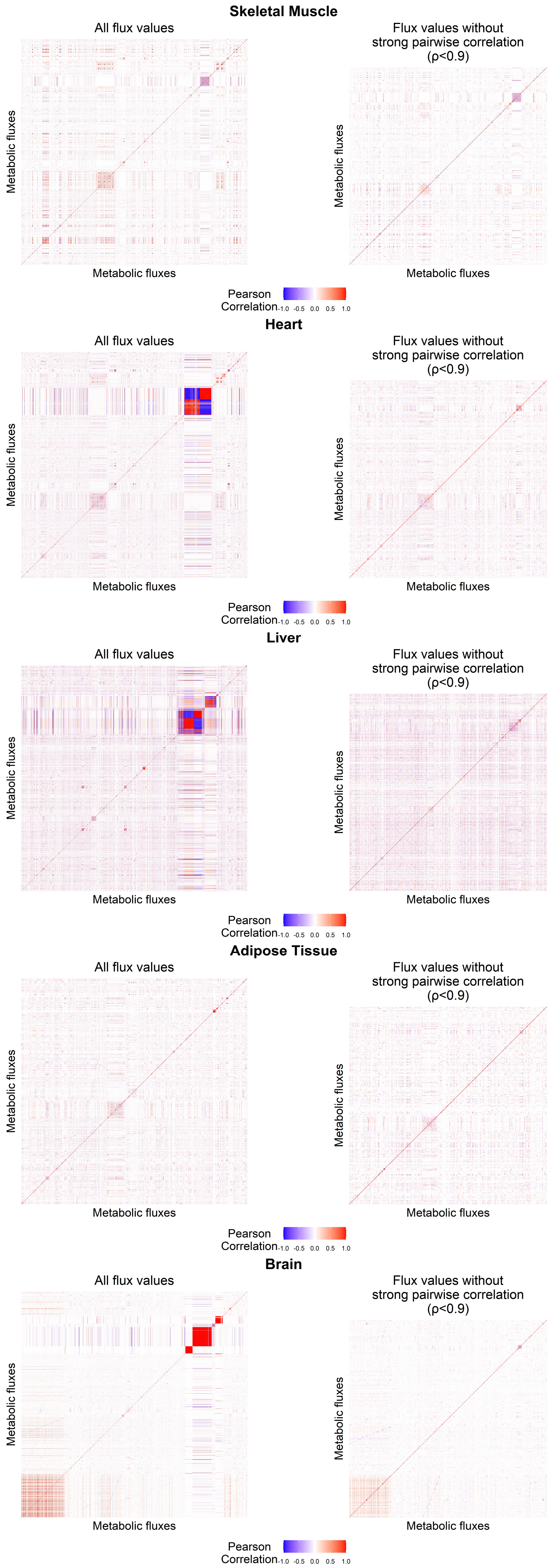
Skeletal Muscle
All flux values
Flux values without
strong pairwise correlation
(ρ<0.9)
Metabolic fluxes
Metabolic fluxes
Metabolic fluxes
Metabolic fluxes
Pearson
Correlation
-1.0 -0.5 0.0 0.5 1.0
Heart
All flux values
Flux values without
strong pairwise correlation
(ρ<0.9)
Metabolic fluxes
Metabolic fluxes
Metabolic fluxes
Metabolic fluxes
Pearson
Correlation
-1.0 -0.5 0.0 0.5 1.0
Liver
All flux values
Flux values without
strong pairwise correlation
(ρ<0.9)
Metabolic fluxes
Metabolic fluxes
Metabolic fluxes
Metabolic fluxes
Pearson
Correlation
-1.0 -0.5 0.0 0.5 1.0
Adipose Tissue
All flux values
Flux values without
strong pairwise correlation
(ρ<0.9)
Metabolic fluxes
Metabolic fluxes
Metabolic fluxes
Metabolic fluxes
Pearson
Correlation
-1.0 -0.5 0.0 0.5 1.0
Brain
All flux values
Flux values without
strong pairwise correlation
(ρ<0.9)
Metabolic fluxes
Metabolic fluxes
Metabolic fluxes
Metabolic fluxes
Pearson
Correlation
-1.0 -0.5 0.0 0.5 1.0

**Fig. S4: Heatmaps of pairwise Pearson correlation coefficients between reaction flux values before and after filtering out reactions with strong pairwise correlations**. Reaction fluxes in each organ are ordered based on hierarchical clustering of correlation coefficients. To remove reaction flux pairs with strong correlation, for each pair of reaction flux values with $\rho>0.9$ the feature with the largest mean absolute correlation to other flux values was removed.

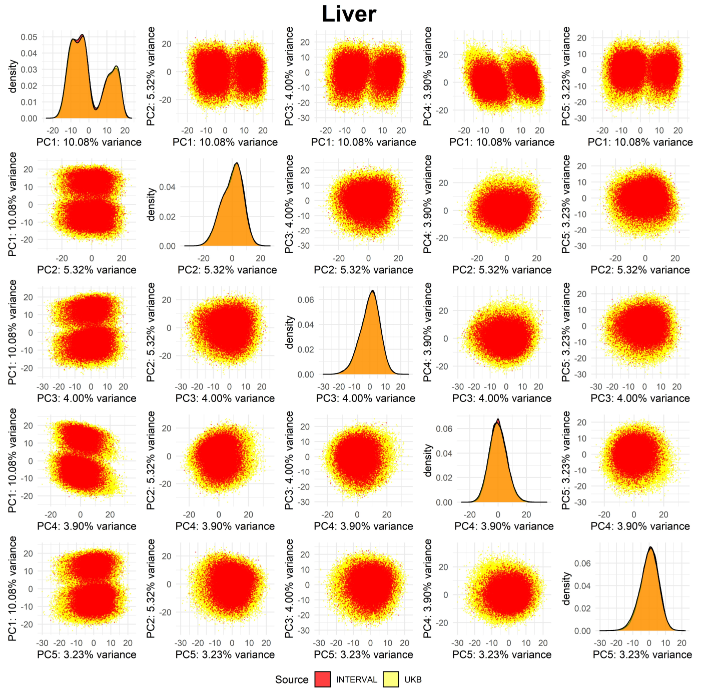
Liver
density
PC1: 10.08% variance
PC2: 5.32% variance
PC3: 4.00% variance
PC4: 3.90% variance
PC5: 3.23% variance
Source INTERVAL UKB

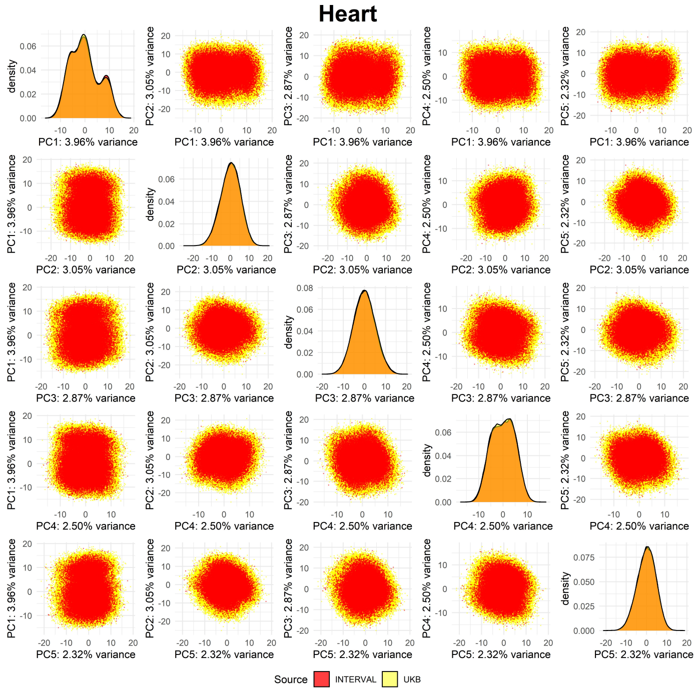
Heart
density
PC1: 3.96% variance
PC2: 3.05% variance
PC3: 2.87% variance
PC4: 2.50% variance
PC5: 2.32% variance
Source INTERVAL UKB

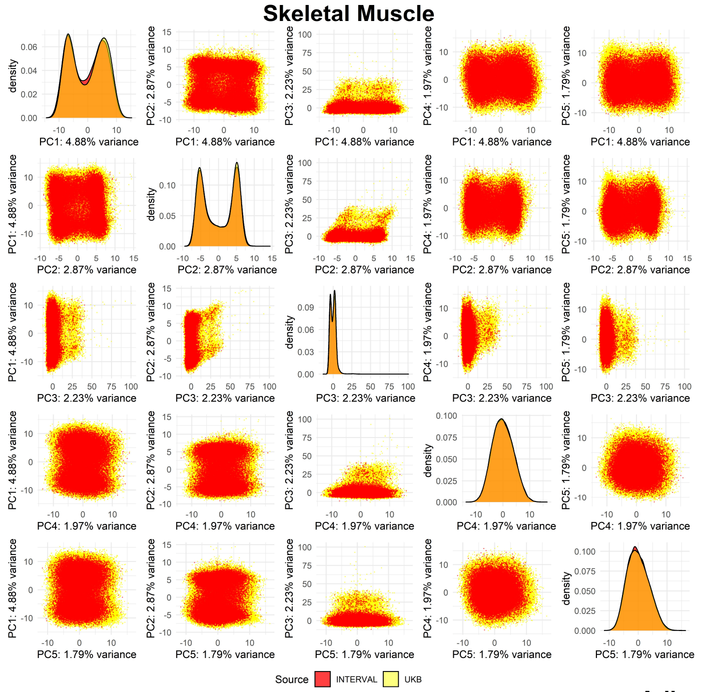
Skeletal Muscle
density
PC1: 4.88% variance
PC2: 2.87% variance
PC3: 2.23% variance
PC4: 1.97% variance
PC5: 1.79% variance
Source INTERVAL UKB

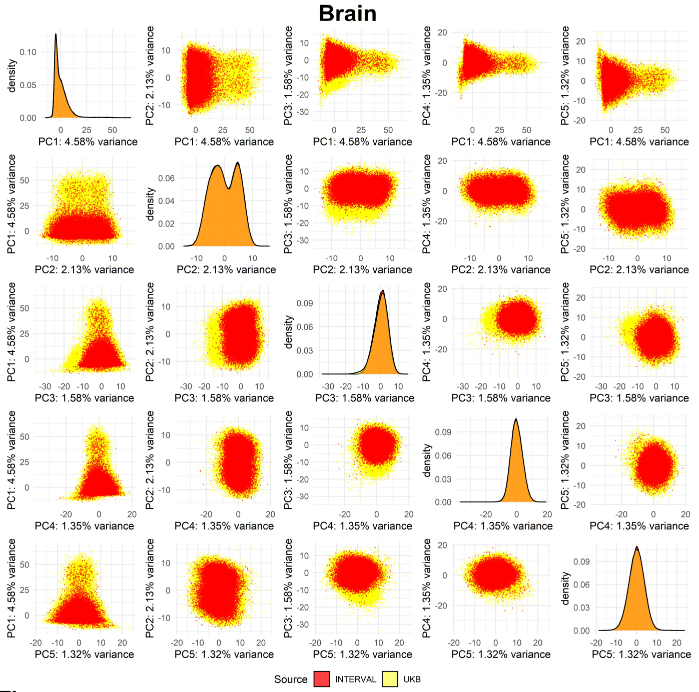
Brain
density
PC1: 4.58% variance
PC2: 2.13% variance
PC3: 1.58% variance
PC4: 1.35% variance
PC5: 1.32% variance
Source INTERVAL UKB

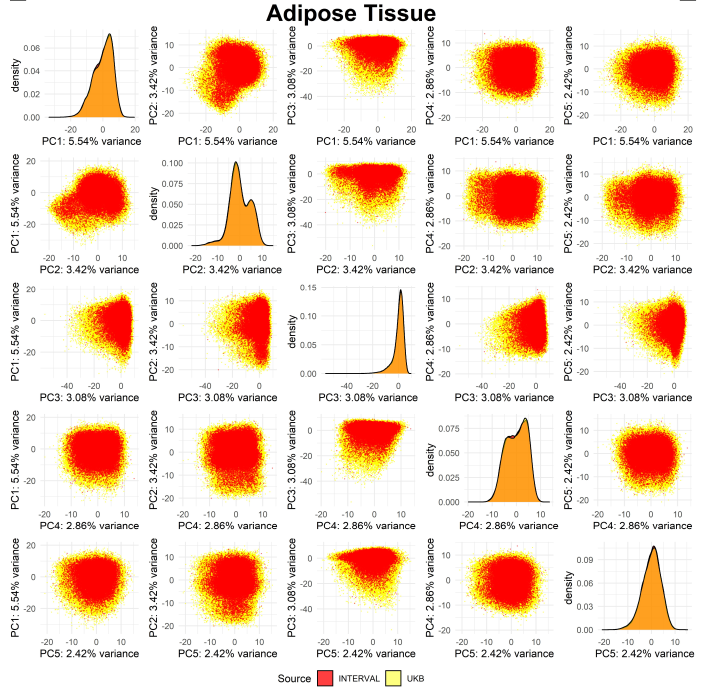
Adipose Tissue
density
PC1: 5.54% variance
PC2: 3.42% variance
PC3: 3.08% variance
PC4: 2.86% variance
PC5: 2.42% variance
Source INTERVAL UKB

**Fig. S5: Principal component analysis**. Pairwise score plots between the first five principal components in UKB and INTERVAL computed using genetically personalised metabolic flux values as features. Each organ is analysed and plotted independently.

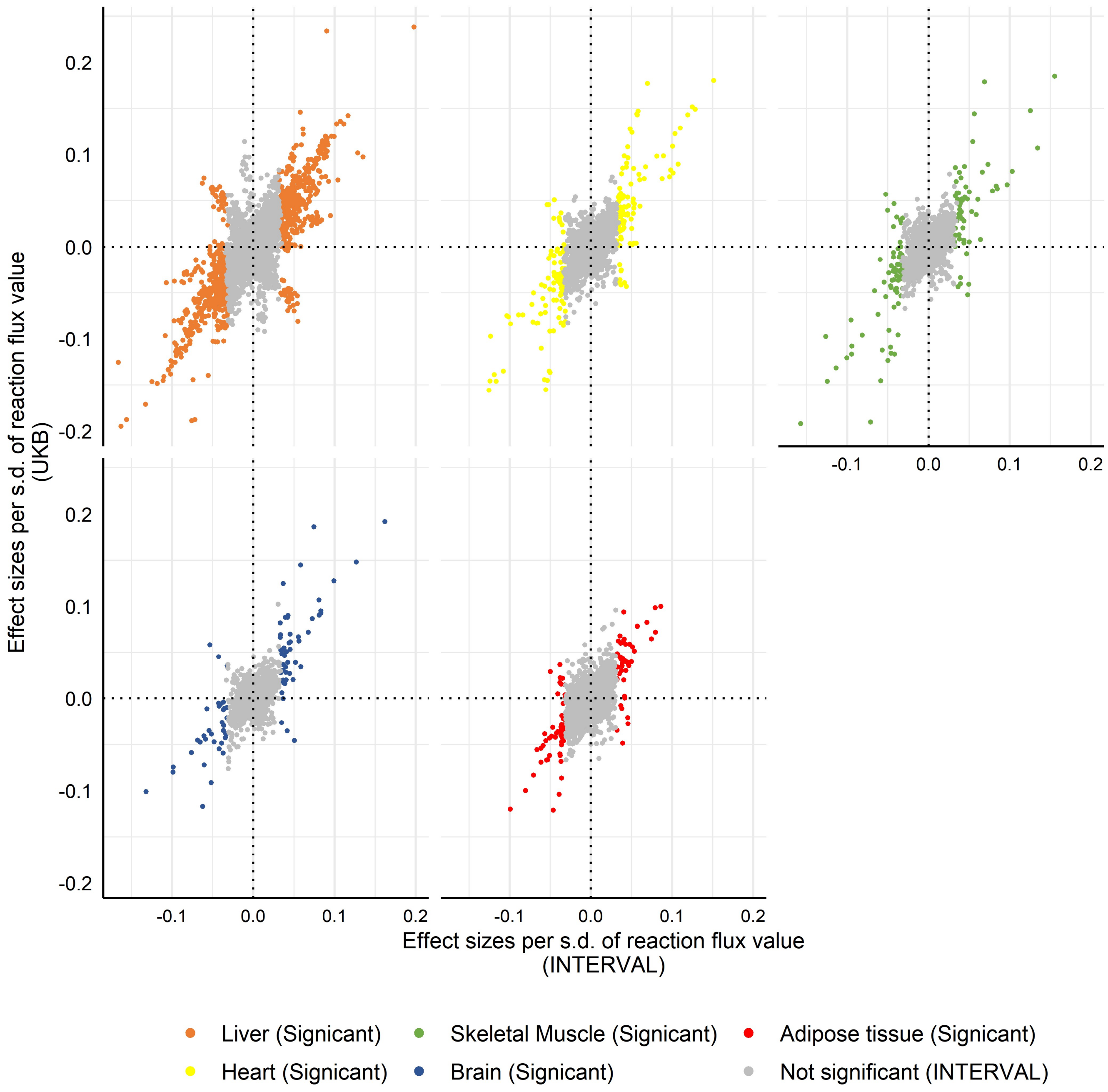


**Fig. S6: Plot of flux effect sizes per organ to blood metabolic features measured with the Nightingale Health platform in INTERVAL and UKB.**

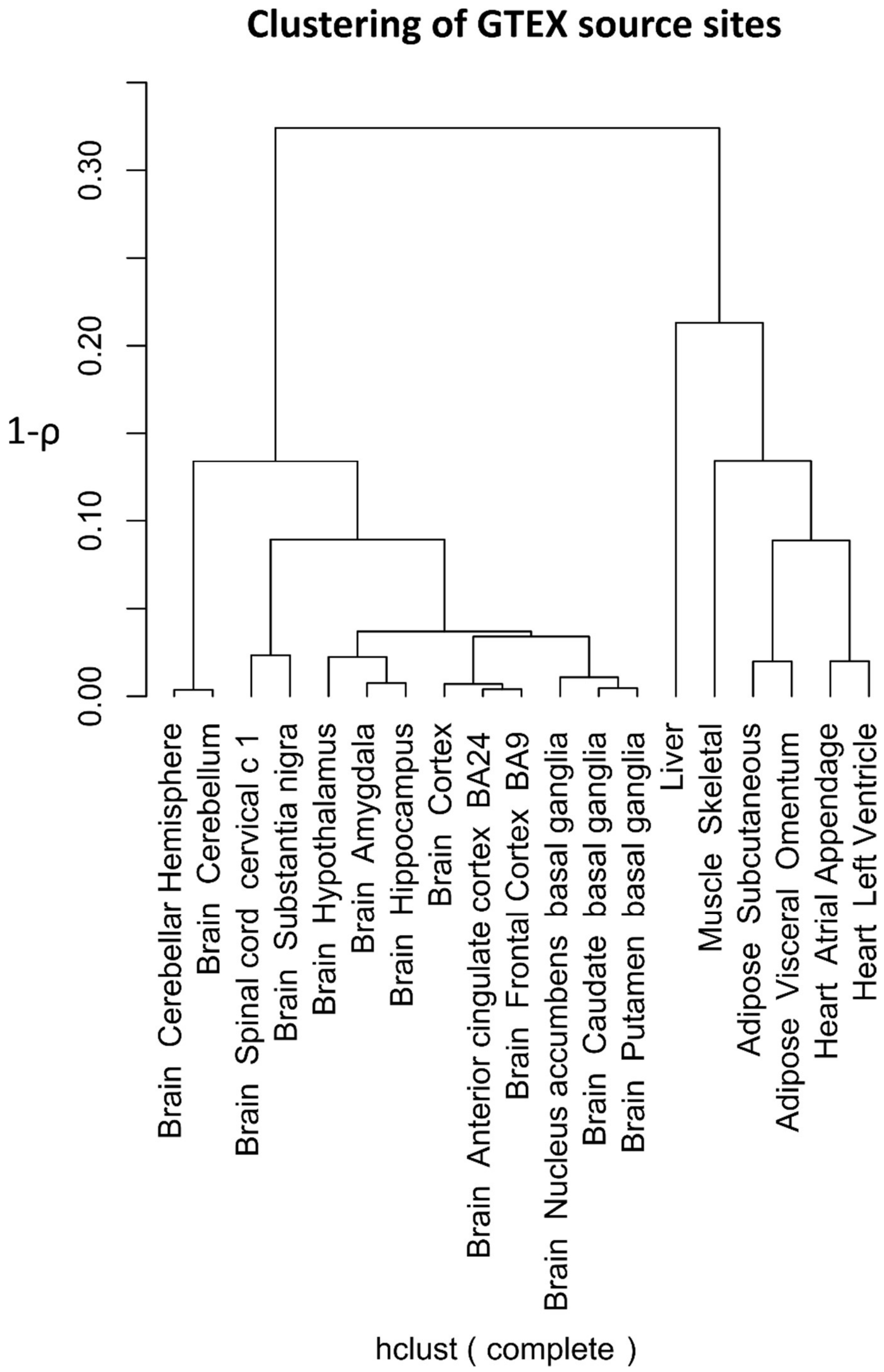


**Fig. S7. Clustering of GTEX source sites.** Source sites are clustered hierarchically using the correlation between average transcripts abundance in each site as a measure of similarity.
